# A Case Study on Colposcopy-Based Cervical Cancer Staging Reveals an Alarming Lack of Data Sharing Hindering the Adoption of Machine Learning in Clinical Practice

**DOI:** 10.1101/2025.08.14.25333566

**Authors:** Maxi Schulz, Andreas Leha

## Abstract

**Background:** The inbuilt ability to adapt existing models to new applications has been one of the key drivers of the success of deep learning models. Thereby, sharing trained models is crucial for their adaptation to different populations and domains. Not sharing models prohibits validation and potentially following translation into clinical practice, and hinders scientific progress. In this paper we examine the current state of data and model sharing in the medical field using cervical cancer staging on colposcopy images as a case example.

**Methods:** We conducted a comprehensive literature search in PubMed to identify studies employing machine learning techniques in the analysis of colposcopy images. For studies where raw data was not directly accessible, we systematically inquired about accessing the pre-trained model weights and/or raw colposcopy image data by contacting the authors using various channels.

**Results:** We included 46 studies and one publicly available dataset in our study. We retrieved data of the latter and inquired about data access for the 46 studies by contacting a total of 92 authors. We received 15 responses related to 14 studies (30%). The remaining 32 studies remained unresponsive (70%). Of the 15 responses received, two responses redirected our inquiry to other authors, two responses were initially pending, and 11 declined data sharing. Despite our follow-up efforts on all responses received, none of the inquiries led to actual data sharing (0%). The only available data source remained the publicly available dataset.

**Conclusions:** Despite the long-standing demands for reproducible research and efforts to incentivize data sharing, such as the requirement of data availability statements, our case study reveals a persistent lack of data sharing culture. Reasons identified in this case study include a lack of resources to provide the data, data privacy concerns, ongoing trial registrations and low response rates to inquiries. Potential routes for improvement could include comprehensive data availability statements required by journals, data preparation and deposition in a repository as part of the publication process, an automatic maximal embargo time after which data will become openly accessible and data sharing rules set by funders.

## 1 Background

The rapid advancements in artificial intelligence (AI) and machine learning (ML) have led to a proliferation of successful applications in various medical domains. Notable examples include text processing for medical record analysis [1], literature understanding (https://elicit.com/welcome, last accessed October 1, 2024), predictive modelling for outcome prediction [2, 3], and image recognition [4]. The metaphor of ’standing on the shoulders of giants’ popularized by Isaac Newton [5] is particularly explicit in ML applications where new applications leverage not only knowledge and ideas of previous studies, but also pre-trained models that can be fine-tuned for specific use-cases by means of transfer learning [6, Chapter 11]. For instance, image classification models in radiology frequently build upon general image recognition models such as ImageNet or ResNet [7]. Similarly, large language models (LLMs) are commonly fine-tuned for medical applications [8–11].

Besides the invention of convolution and the availability of greater compute resources, the inbuilt ability to adapt existing models to new applications has been one of the key drivers of the success of deep learning (DL) models. Thereby, sharing trained models enables the validation of these models both in similar and in different populations and is therefore crucial for their adaptation to different populations and domains [12]. However, missing model validation has been identified as one of the key factors hindering the more widespread application of ML models in clinical practice [13, 14].

The importance of model sharing has been recognised by the field and repositories facilitating model sharing have been created (e.g. Kipoi: [15], Huggingface: https://huggingface.co/, last accessed October 1, 2024). In recent years, foundation models have been published for a lot of applications (Examples listed by application include: Clinical Large Language Models: [10], Single-cell transcriptomics: [16], Medical Image Analysis: [17]) with the explicit intention that they are usable especially as foundation for fine-tuned models in specific applications.

Not sharing models, on the other hand, prohibits validation and potentially following application, thus, translation into clinical practice, and hinders scientific progress by not providing the shoulders for others to stand on.

In this paper we draw on our experiences from cervical cancer staging on colposcopy images to examine the current state of model sharing in the medical field. We discuss the implications of limited model sharing and explore potential routes for improvement, highlighting the need for increased collaboration and data sharing to accelerate scientific progress and translation into clinical practice.

## 2 Methods

Given the absence of a specific reporting guideline for this type of review, our methodology is adapted to the study’s objectives. While this study does not claim to be exhaustive, as only a single database was queried and screening and data extraction was performed by a single reviewer only, we closely followed best practices for systematic reviews.

### 2.1 Use Case: Using DL to classify cervical cancer based on colposcopy images

Cervical cancer causes hundreds of thousands of deaths each year world-wide and has been reported as the leading cause of cancer-related death in women in most parts of Africa [18]. Invasive cervical cancers are usually preceded by a years-long phase of non-invasive abnormalities in cervical tissue [19, Background]. As persistent infections with human papillomavirus (HPV) are known risk factors for cervical cancer, HPV vaccination and HPV-based screening for percursor lesions are suggested strategies for cervical cancer prevention [20]. During the phase of transformation from precursor lesion to invasive cancer, differentiation between cervical cancer and different states of pre-invasive abnormalities is important for treatment stratification from low frequency follow-up to high-frequency follow-up or immediate treatment via ablation or excision. Cervical intraepithelial neoplasias (CIN) can be graded according to characteristics such as the proportion of non-differentiated cells and nuclear abnormalities [19, Annex 4, Cancer and pre-cancer classification systems]. Colposcopy offers an accurate way to diagnose and grade CIN [21]. As an image based technique colposcopy lends itself towards application of DL models, which have proved to be especially powerful for image analysis [22].

An initial literature search revealed that, indeed, DL models for classification of the progression grade of cervical cancer had already been trained on very large datasets from colposcopy and some even had been evaluated in external populations. Many of these existing studies offered data sharing upon request. In order to validate the performance on the local population and/or to yield a model fine-tuned to the local population we subsequently tried contacting the existing studies for access to the pre-trained model weights and/or to the datasets.

### 2.2 Literature search

We conducted a comprehensive literature search in PubMed to identify studies employing ML techniques in the analysis of colposcopy images. As primary studies we included studies for data inquiry that met the following criteria: application of ML methods using images from colposcopy with the intent to support treatment decisions for cervical carcinoma, development and/or validation studies, classification and/or segmentation models. We excluded studies that met any of the following criteria: systematic reviews and/or overviews of existing literature in the field, classification models with ground truth based on expert opinion rather than biopsy, classification tasks unrelated to CIN classification (e.g., quality classification of the cervical image), or too few data (fewer than 200 images).

We further identified studies and data sources for ML-based analysis of colposcopy images referenced by the primary studies, which may compromise studies and online resources. The date of the last search was 5 May 2024. In Appendix A.1 and A.2 we provide a detailed description of the search strategy and selection process for included studies.

### 2.3 Data availability

We categorized the included studies and online resources into two groups: analysis publications (studies only) and publications that provide data (i.e. studies or online resources referenced as data sources for training or validation of ML-based models in one or multiple primary studies). For analysis publications, we extracted information on the data used and further distinguished between those that employed their own dataset and those that utilized external data sources.

We assessed the presence of a data availability statement (DAS) for each included study. When a DAS was present, we extracted its content and classified it into two types: generic and specific. A generic DAS lacks specificity regarding the type of available data, whereas a specific DAS provides details on the type of data available and the access procedure. For studies with specific DAS, we further categorized the information provided into three subcategories: (1) availability of raw data, (2) availability of training code, and (3) availability of pre-trained model weights. We also noted the access procedures for each of these categories, where applicable.

We further categorized the DAS into two types: positive DAS and negative DAS. A positive DAS is defined as a statement that explicitly mentions the availability of any of the three categories (raw data, source code, or pre-trained weights) or generally refers to available “data”. This may also include statements where data can be inquired upon request. In contrast, a negative DAS is defined as a statement that declines data sharing for any of the three categories or generally refers to the unavailability of data.

In Appendix A.3 we provide further details on data extraction from the included studies.

### 2.4 Data inquiry

Where data was directly accessible, we accessed the data via download and assessed the usability of the data for our model training in terms of outcome definition and number of cases.

Where data was not directly accessible, we systematically inquired about accessing the pre-trained model weights and/or raw colposcopy image data for the purpose of our training. For publications that focussed on providing data and for analysis publications that evaluated previously trained ML-based models, we inquired about accessing the datasets only. For analysis publications with a negative DAS (i.e. data is declared unavailable) we inquired about accessing the pre-trained model weights only. In all other cases, we inquired about accessing both the pre-trained model weights and the raw colposcopy image data.

We initiated the inquiry by contacting the corresponding author(s) or individuals listed as contact persons in the DAS through email. If we did not receive a response within two weeks of the initial contact, we dispatched a follow-up email. For Chinese researchers, we ensured at least one of these two emails were sent from a secondary email address to lower the chances of our emails from being blocked by filters.

If contact was not established in the next two weeks, we then reached out to both the corresponding author(s) and the first author (if different) using the academic networking site, ResearchGate, via messaging, where accounts were available. In situations where email addresses were no longer valid, we contacted the authors directly using ResearchGate.

For each step of the data inquiry process, we recorded the number of contacted authors and corresponding studies that responded, that did not respond and those that could not be reached (due to invalid email addresses or missing ResearchGate accounts). If an author was contacted for multiple studies, their response was applied uniformly across all corresponding studies.

We classified the responses to our data inquiry requests into four categories: (1) data sharing declined, where the author(s) declined to share data; (2) data sharing successful, where the author(s) provided data and facilitated its upload; (3) pending response, where we awaited further communication or clarification from the author(s); and (4) redirect/referral, where the author(s) redirected our inquiry to an alternative contact person. For each decline, we documented the reason(s) stated by the author. We addressed each received response by a follow-up email. We suggested alternative ways for data sharing for data sharing declines (1), provided a link for the anonymized upload of model weights and/or raw data to our locally hosted servers (2), reminded author(s) through a follow-up email (3) and contacted the alternative contact person(s) (4).

If we received no response to our initial email or ResearchGate inquiry, no response to our follow-up email and/or contact attempts failed, we concluded that data sharing was not feasible for that particular study. The last date of registering responses to our inquiries was September 30, 2024.

### 2.5 Response Analysis

We summarized and aggregated the number of responses per study as well as the data sharing outcome per study. We explored potential associations between study characteristics and response receipt (no response / response) and data sharing outcomes (data shared / data not shared). Specifically, we tested the following study characteristics for association:

- Status of the DAS (DAS included, DAS not included)
- Content of the DAS (Positive, negative)
- Region of publication origin (Country of first author categorized by continent)
- Time of publication (year)
- Size of the study (in total number of image data used)
- Successful model training in the study (performance metrics of the best-performing model on test or validation dataset, including Area Under the Curve (AUC) and Accuracy (only applicable for classification models))

We performed a chi-square test of independence with p-values simulated by Monte Carlo simulation with 2,000 replications to examine the independence of response receipt and data sharing outcome from nominal variables, and logistic regression analysis to investigate the relationship between continuous variables and response receipt and data sharing outcome. A significance level of *α* = 0.05 was assumed.

## 3 Results

### 3.1 Included studies

After removing a duplicate preprint entry, the literature search yielded 276 records for screening. Of these, 199 records were excluded and 5 of these articles could not be retrieved, leaving 72 articles for full-text screening. Of these, 38 studies met our inclusion criteria. Additionally, we retrieved and screened 9 records from citations of the screened literature which compromised 8 studies and one internet resource that provided a publicly available dataset of colposcopy images. In total, 46 studies and one publicly available dataset were included for data inquisition (see Supplement 2 for a flow chart of the study inclusion process and Appendix C for a list of included studies).

### 3.2 Characteristics and data availability of included studies

The 47 included records comprised 39 studies that performed ML-based analysis on colposcopy images (analysis publications) and 8 records that provided data sources, including one publicly available dataset and seven studies (publications providing data).

Of the 39 analysis studies, 7 used external data sources, with some publications using multiple external data sources. Colposcopy images from the Guanacaste project were most often used as external data source (5), followed by the ALTS study (4) and the Costa Rica Vaccine Trail (CVT) (3). Further, data from the Kaggle Dataset (publicly available dataset) (2), DYSIS (1), the Biopsy study (Biop) (1) and Biopsy Study - Europe (D Biop) (1) was used (refer to Table B1 for more information on the data sources). The other 32 studies used their own datasets or did not report on using external data sources explicitly.

The data for the publicly available dataset was directly accessible via the Kaggle platform as part of the “Intel & MobileODT Cervical Cancer Screening Competition” [23].

Of the 46 studies, 24 provided data availability statements (DAS), while 22 did not (see Figure 1 A). Of note, none of the 7 publications that explicitly provide data, of which six were used as external data sources in one or multiple of the primary studies (see above), included a DAS. Out of the 24 publications that did include a DAS, 20 relied on their own datasets, while four utilized external data sources.

**Fig. 1.**
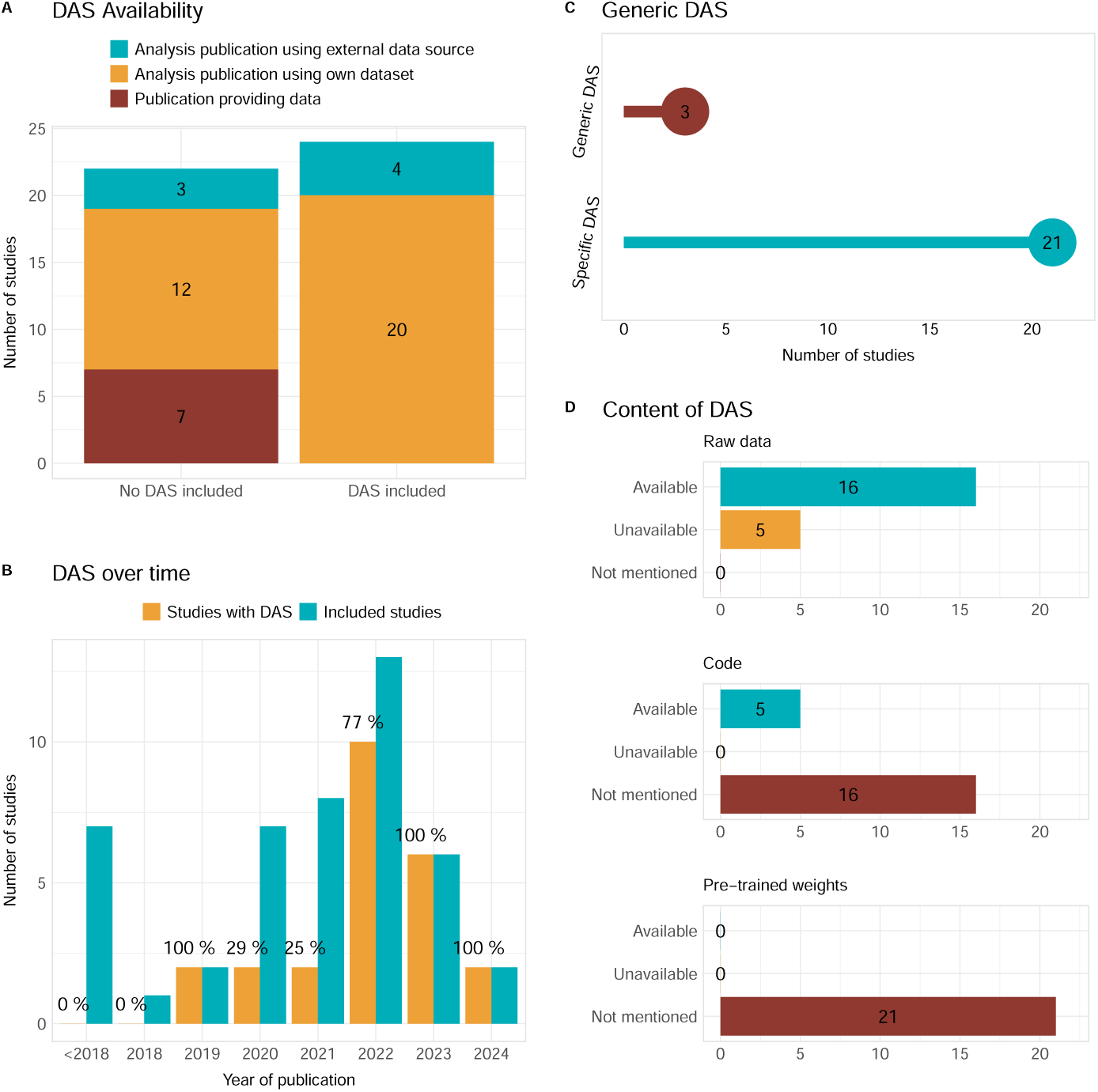
Status and content of the DAS of included studies (excluding the publicly available dataset).

The inclusion of a DAS in publications is a reflection of evolving publishing practices over time. In our study, we observe that none of the works published up until 2018 provided a DAS (see Figure 1 B). However, starting from 2019, there is a gradual increase in the proportion of studies featuring a DAS. For studies from 2020/2021, about 25% included a DAS. This noticeably rose to 77% in 2022 and reached 100% in both 2023 and 2024.

Among the 24 studies that provide a DAS, three include a generic DAS compared to 21 studies with a DAS specifying what type of data is available and how it can be accessed. 16 of these 21 studies declare their raw data available, with all of them accessible upon reasonable request from the corresponding author(s). This compares to 5 studies that declare their raw data unavailable. Furthermore, 5 studies make their source code directly available through a link to an external repository, whereas 16 do not mention the availability of the source code. Neither do any of the 21 studies mention the availability of pre-trained weights. (Figure 1 D) In total, the three studies with a generic and 18 of the 21 studies with a specific DAS provide a positive DAS, declaring any of the three categories (raw data, source code, pre-trained weights) available, compared to three studies that provide a negative DAS.

### 3.3 Data sharing outcomes and responses

We accessed the publicly available dataset of colposcopy images via login to the Kaggle platform. This dataset consists of 8,727 images of the cervix, captured using a smartphone-based digital cervical imaging device. Each image is annotated with a cervix type label, which identifies the location of the transformation zone. We determined that this dataset is suitable for training a model to identify the cervix type, which could be a useful component of a CIN classification algorithm. However, we concluded that the dataset alone is insufficient for training a reliable CIN classification algorithm.

Of the remaining 46 included studies, we attempted to contact the authors for access to datasets, pre-trained models, or both. We requested access to datasets only for 10 studies, datasets along with pre-trained models for 31 studies, and pre-trained models only for 5 studies.

#### 3.3.1 Initial Email Inquiry

We initiated our data access inquiry by emailing 68 authors, comprising 54 unique individuals, associated with the 46 studies (see Figure 2). Note that some individuals were contacted more than once due to their inclusion as both corresponding and first authors across multiple studies. For the purpose of this report, we will present all numbers based on the total number of authors contacted, rather than unique individuals. We contacted corresponding authors (38 emails), first authors (4 emails), authors with both roles (18 emails), and other authors (8 emails). Note that first authors were only contacted if they were explicitly mentioned in the DAS or otherwise referenced in relation to data access.

**Fig. 2.**
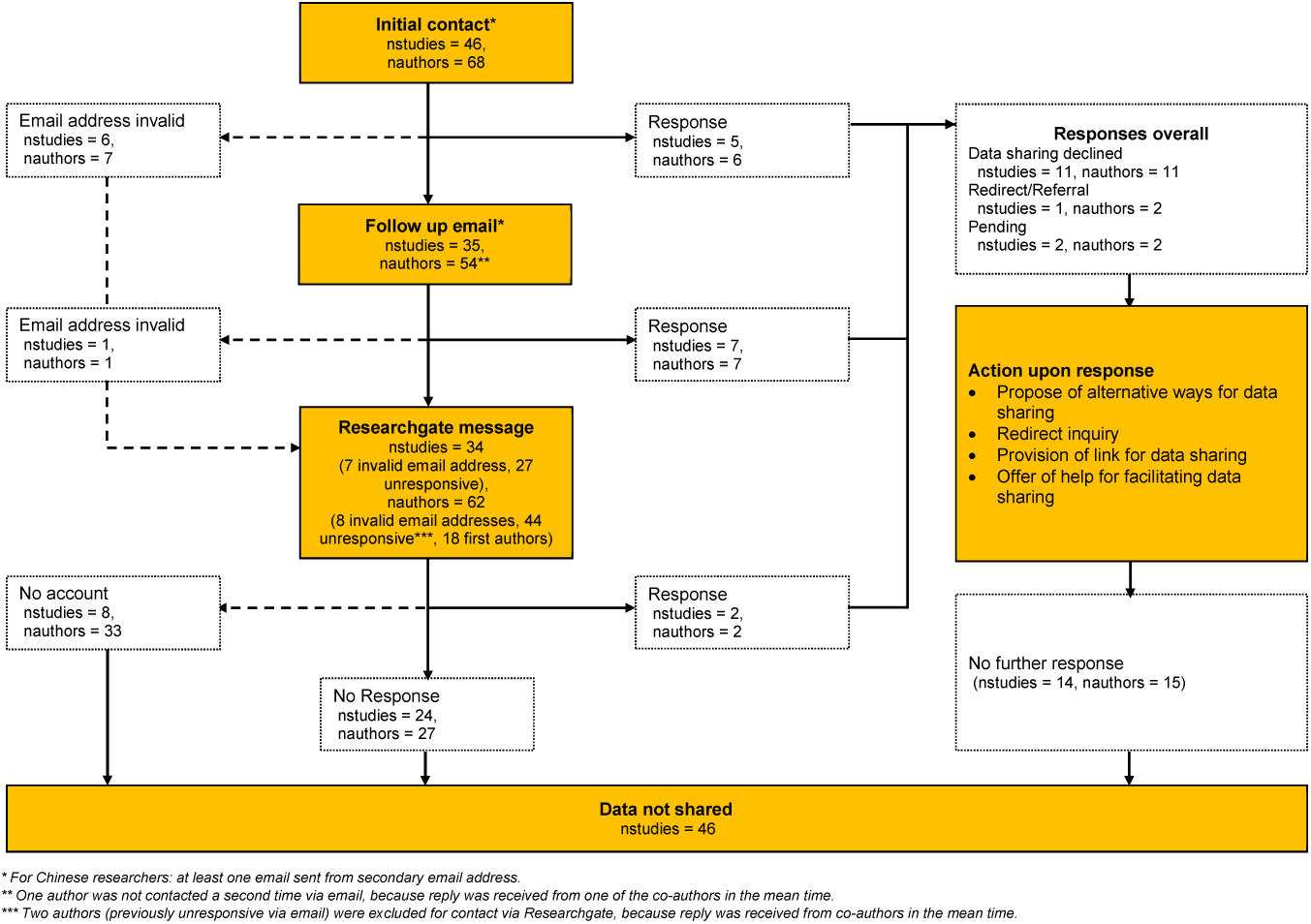
Responses and data sharing outcomes following the data inquiry process.

We encountered issues with 7 authors’ email addresses, which were no longer valid. We received 6 responses from 3 unique authors, while the majority of email attempts went unanswered with 55 out of 68 attempts.

Following the first round of inquiries, we were unable to get a response for six studies due to invalid email addresses. We received responses related to five studies, while 35 studies remained unresponsive.

#### 3.3.2 Follow-up Email Inquiry

We sent a second email inquiry to the authors of the 35 studies that had not responded to our initial contact. This round involved 54 contact attempts (one previously contacted author was not contacted a second time because we received a reply from a co-author), targeting corresponding authors (35 emails), first authors (4 emails), authors with both roles (11 emails), and other authors (4 emails).

One email address was found to be invalid, affecting one study. Of the remaining 53 attempts, we received 7 responses from 4 unique authors. The majority of our attempts (46 out of 54) again went unanswered.

After the second round of email inquiry, 27 studies still had no response, while we had received responses for 7 studies, and one study was affected by an invalid email address.

#### 3.3.3 ResearchGate inquiry

For the 27 studies that remained unresponsive, as well as the 7 studies with invalid email addresses, we attempted to contact the authors through ResearchGate as an alternative mean. Specifically, we reached out to the previously contacted authors, as well as the first authors who had not been contacted via email. We identified 44 authors who had been previously contacted without success, with 21 having a ResearchGate account. We also identified 18 first authors who had not been previously contacted via email, with 8 having a ResearchGate account. In total, 62 authors were eligible for inquiry via ResearchGate, with 29 having an account. We proceeded to inquire with all authors who had a ResearchGate account through the platform. Due to unavailable accounts, 8 studies out of 34 could not be reached via ResearchGate.

We received two responses from one unique author, corresponding to two studies, while the other 27 attempts remained unresponsive.

On a study-level, we received responses for 2 studies, while 24 remained unresponded.

#### 3.3.4 Content of responses

We received 15 responses from a total of 92 contacted authors (through email and/or ResearchGate message) across the 46 studies, with 8 unique authors accounting for multiple studies. We received two responses for one study, therefore the 15 responses corresponded to 14 studies. The remaining 32 studies remained unresponsive (including the studies for which contact failed due to invalid email addresses and/or missing ResearchGate accounts).

Of the 15 responses received, two responses redirected our inquiry to other authors for further information on data sharing. We subsequently contacted these authors following the data inquiry process.

Two responses were pending, awaiting further communication or clarification. In one case, the authors expressed willingness to share data but required further clarification on the matter. We thanked the author and offered our assistance in facilitating data sharing. In another case, the authors responded that they were willing to share their data. We provided a link for upload of the data. In both cases we followed upon these responses by follow up emails, however, we did not receive any further response or data via upload.

The remaining 11 responses indicated that data sharing would not be feasible, i.e. data sharing was declined. The authors cited various reasons for not sharing data, including lack of resources and funding (4 responses) and confidentiality agreements and privacy agreements with hospitals (5 responses). Two responses did not provide a reason. Notably, for three studies with a privacy agreement with the hospital, an additional reason for not sharing data was the ongoing trial registration of the tool (response of one author accounting for multiple studies). For all declines, we sent follow-up emails suggesting alternative ways of data sharing and/or stating that we would be open to further collaboration if data sharing became feasible at some point in the future.

Despite our follow-up efforts on all responses received, none of these follow-up emails received a response, and none of the inquiries ultimately led to actual data sharing (see Figure 2 and Table 1). As a result, the only available data source remained the publicly available dataset.

**Table 1.** Number of contacted authors, responses received and data sharing outcomes. In Brackets: The number of corresponding studies.

|  | Contacted authors<br>(Contacted publications) | Responses<br>(Responsive publications) | Data shared |
| --- | --- | --- | --- |
| Initial Email | 68 (46) | 6 (5) | 0 |
| Follow-up Email | 54 (35) | 7 (7) | 0 |
| ResearchGate | 29 (26) | 2 (2) | 0 |

### 3.4 Factors influencing data sharing and response behavior

Figure 3.4 presents the number of studies by data sharing outcome and responses received, categorized by the status of their DAS. Notably, none of the 46 studies that were eligible for data inquiry shared their data, regardless of the status of their corresponding DAS. We received responses from a total of 14 studies, which were further categorized as follows: 5 studies did not include a DAS, one had a negative DAS, and 8 had a positive DAS. In contrast, we received no response to our inquiries from 32 studies, which were categorized as follows: 17 studies did not include a DAS, two had a negative DAS, and 13 had a positive DAS. Given that none of the contacted studies provided data sharing, we omitted the analysis of data sharing outcomes.

**Fig. 3.**
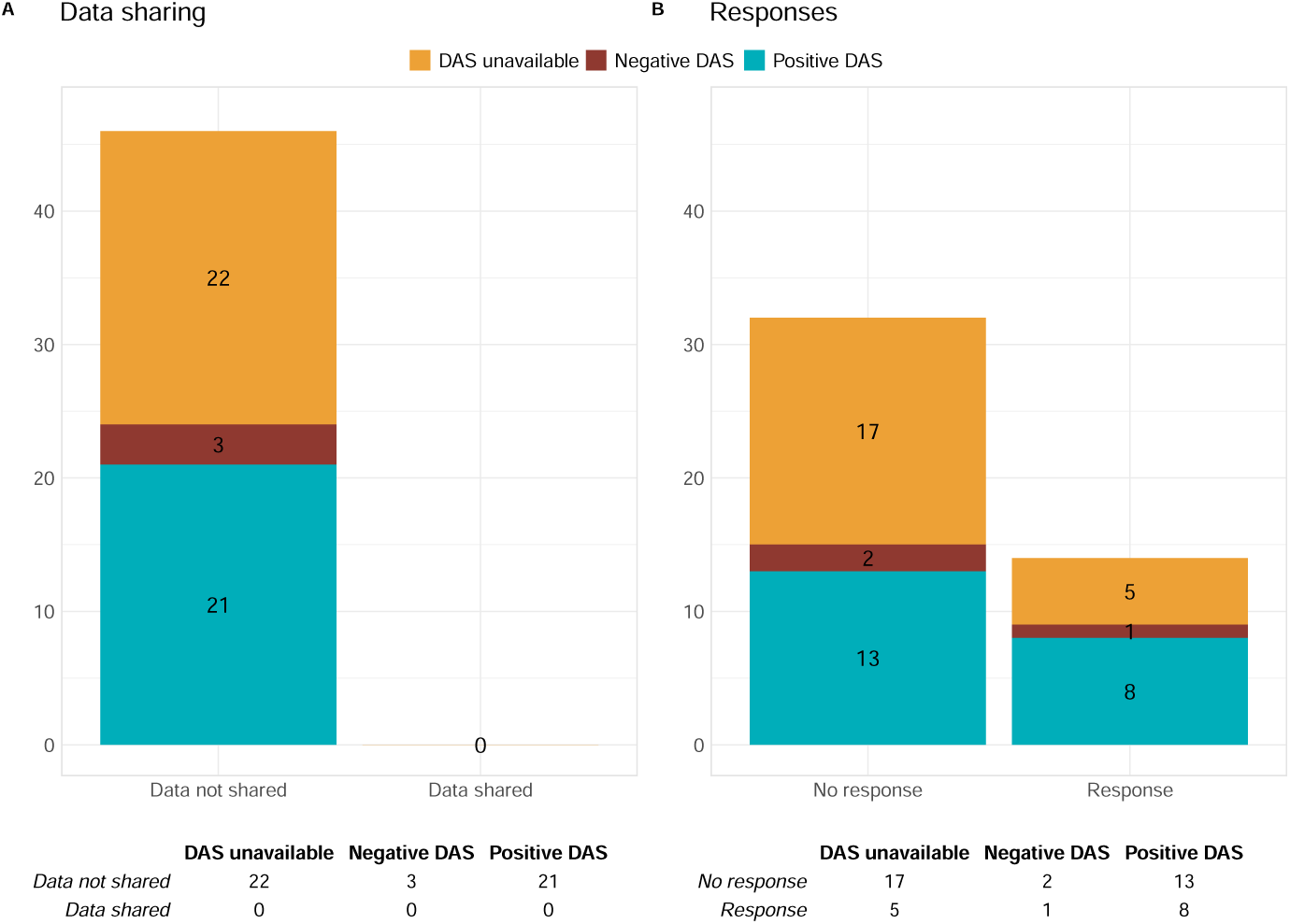
Number of studies by data sharing and responses received

The results of our analysis on the factors influencing response behavior (on study-level) are presented in Table 2. The analysis showed that none of the investigated factors proved a statistically significant association with response receipt. Most notably, our analysis did not reveal a statistically significant association between the status of the DAS and the response behavior, *χ*^2^ (N=46) = 1.21, *p* = .697.

**Table 2.** Results for tests of association between study characteristics and response behavior.

| | | Frequencies<sup>1</sup> | | $\chi^2$ | *P*-value<sup>2</sup> |
| --- | --- | --- | --- | --- | --- |
|  |  | No response | Response |  |  |
| Status<br>DAS | DAS unavailable | 17 (53%) | 5 (36%) | 1.183 | 0.335 |
|  | DAS available | 15 (47%) | 9 (64%) |  |  |
| Content<br>DAS | Negative DAS | 2 (13%) | 1 (11%) | 0.025 | 1 |
|  | Positive DAS | 13 (87%) | 8 (89%) |  |  |
| Region | Asia | 20 (63%) | 9 (64%) | 1.502 | .679 |
|  | Europe | 3 (9%) | 0 |  |  |
|  | North America | 9 (28%) | 5 (36%) |  |  |
| | | Mean ± Std deviation | | Z<br>statistic | *P*-value<sup>3</sup> |
|  |  | No response | Response |  |  |
| Year<sup>4</sup> | | 4.56 ± 5.41 | 4.07 ± 5.06 | -0.294 | .769 |
| Number of image data | | 6,686 ± 8,775 | 14,430 ± 25,341 | 1.221 | .222 |
| AUC<sup>5</sup> | | 0.86 ± 0.1 | 0.9 ± 0.07 | 1.040 | .298 |
| Accuracy<sup>6</sup> | | 0.78 ± 0.17 | 0.85 ± 0.06 | 0.992 | .321 |

<sup>1</sup>Percentages are reported column-wise.
<sup>2</sup>Based on $\chi^2$ test of independence with *p*-values computed by Monte Carlo simulation with 2,000 replications.
<sup>3</sup>Based on logistic regression analysis.
<sup>4</sup>Defined as the difference in years from 2024.
<sup>5,6</sup>Numbers are reported for studies that included AUC and/or Accuracy metrics. The best metric per study is used. Number of studies who reported AUC metrics: 22, Number of studies who reported accuracy metrics: 21.

## 4 Discussion

With the aim of a locally fine-tuned model, we conducted a systematic literature search for studies on staging of potential cervical cancer patients based on ML-based analysis of colposcopy images. Our search yielded 46 studies that had trained models or presented training data, as well as one additional publicly available dataset. While the publicly available dataset offered accessible data, it was insufficient on its own to train a reliable CIN classification model. We subsequently contacted the authors of the remaining 46 studies for data access, but none of our inquiries resulted in actual data sharing. The amount of existing studies suggests that model validation and transfer learning to adapt existing models to the local setting could have been possible. Furthermore, training a new model using one of the high-volume external datasets would have been feasible.

Unfortunately neither pre-trained models nor data sets have been shared upon request. We made several attempts to contact the corresponding and/or first authors through different channels to ask for access to the pre-trained models and/or training data of the 46 studies. For only 14 (30.4%) of the studies we received any answer. The disillusioning result was that we were given access to either the pre-trained model or training data for 0 (0%) of the studies. Thus, the state of data sharing does not seem to have improved from 2009 when 9% of the publications studied by Alsheikh-Ali et al. fully shared their raw data [24].

It is especially discomforting that data sharing was not even possible for datasets that had been shared previously, e.g. colposcopy images from the Guanacaste project which had been used to train and validate DL models several times [25–29], or for publications that explicitly construct and describe datasets aimed at facilitating the training of deep learning models [30, 31]. The MSCI dataset [31] even has a repository on github (https://github.com/yuyao1314/MSCI-dataset, last accessed October 1, 2024) seemingly making the data available, but this repository only contains a few test cases and an email address. Our emails to this address went unanswered.

Some reasons for the gap between the high number of publications on ML applications in the medical domain and the low number of ML models implemented in clinical routine have been proposed: Lack of transparency and poor reporting, high risk of bias and ethical concerns, too optimistic predictive performance, difficulties in interpretation of model decisions and lack of prospective validation [13]. Our systematic analysis of data availability from studies for a specific clinical application revealed that the lack of model sharing poses an additional barrier to the implementation of ML models in clinical practice. Without the availability of pre-trained models, neither validation nor application can be attempted. As a result, locally applicable models need to be trained from scratch, or at least from more general models, since transfer learning or fine-tuning of existing models trained for similar purposes is not possible. To promote data sharing among researchers, it is essential to establish incentives that reward those that “conduct rigorous, transparent and reproducible research” [32]. Despite first findings that indicate a small positive citation advantage (4.3% on average) for publications that make their data available [33], none of the included studies from our case study provided data directly or upon request. One possible approach to incentivize data sharing is the implementation of data sharing policies by journals. Indeed, the past decade has seen an increase in data sharing policies required by journals, with the use of DAS being one method [34]. However, numerous studies have shown that the requirement of a DAS is not sufficient to ensure actual data sharing practices across various fields of applications (Reanalysis of randomized controlled trials: [35], Investigation of data availability in medRxiv preprints: [36], Data availability in urology: [37]). Our analysis confirms these findings, as neither the presence nor the content of a DAS was indicative of receiving a response (p = .335 and p =1) or gaining access to the data. Furthermore, none of the other factors we examined, including publication origin, year of publication, number of image data, and performance metrics, showed an effect on response receipt and data sharing.

From the few responses we received, the stated reasons for not sharing the pretrained model and/or training data included a lack of resources and funding for provision of the data, confidentiality or privacy agreements with hospital partners as well as on ongoing trial registration.

Sharing data can be a time-consuming task: The data or the model may need detailed explanation or scripts for model training which need to be prepared and synchronized for reproducibility of results. A single upload might not suffice. It is our view that sharing data and/or trained models will be more work than not sharing data and/or trained models, and we see only few incentives for authors to invest their time into sharing things with the research community. Furthermore, some authors might even view the data and trained models as their own property. Keeping control over the data and the data sharing has been described as a way to retain power in the scientific system [38]. Given how walled gardens hinder the progress in development of ML models and the application of ML models in clinical practice, we argue that this time needs to be invested and that authors need to be persuaded to give up on their believe of possessing data.

While it is natural to grant any researcher the right to benefit from collected data and the trained model through generating publications from them, we believe that research data and results should be made publicly available at least after some time of embargo. This is especially true for publicly funded research where the public might be rightly called the owner.

How data protection issues also affect the trained models is still in active debate as trained models might leak training instances [39]. Data protection is indeed a strong reason for not sharing data and careful patient consent needs to be obtained. But as the many published data sets in the cancer imaging archive (TCIA) demonstrate, this is possible [40].

We can only speculate about the reasons for not even responding to data requests. Probably time constraints keep answering emails from strangers on data sharing just out of the priority list of authors.

Among the authors contacted via email, we found 8 (11.7%) email addresses failing. Email addresses easily become invalid, for instance, when authors change their institution. Such *email decay* has been recognized as a problem for a long time [41] and poses a challenge for data sharing practices. Consequently, typical data availability statements claiming that “data is shared by the corresponding author upon reasonable request” will become untenable at some point in time.

It is known that the imposter syndrome is present in data scientists [42], so there might also be cases where authors are afraid to fully share their models and or data for fear of potential errors in their work to be easier discovered. This is a serious problem and some change in the attribution of credit needs to happen. A reportedly well performing model that can’t be shared and validated should be less credited than a fully open model, even if there might be errors discovered in that second model.

While there may be cases where data sharing is simply not possible, low response rates to inquiries for data access are a significant barrier for data sharing [37, 43, 44]. Our own experience and that of other researchers highlights that the lack of access to data hinders scientific progress [45]. For instance, Fleetcroft et al. received only 6 responses (20%) from 30 studies, with only one offering potential access to their data [45]. The problem is particularly pronounced for publications with a DAS stating that data is available on request, yet response rates remain low, such as in publications from urology journals where only 28.1% of authors responded to requests for data, although DAS were initially positive [37]. Not only does the DAS itself fail to guarantee adherence to declared data sharing, but it also appears to be limited by a narrow focus on ’raw data’. In our case example, all included studies with a DAS mention the availability of raw data, and some mention and share their source code alongside publication. However, none of the DAS of the included studies explicitly mention the availability of pre-trained weights. While the availability of pre-trained weights may be specific to ML-based publications, the field would benefit from a more comprehensive DAS that requires the mentioning of raw data, source code, and pre-trained weights. Given the limitations of DAS, alternative incentives for data sharing must be explored. Thereby, a collaborative effort among stakeholders, including funders, publishers, societies, institutions, editors, reviewers, and authors, is necessary to create a culture that rewards reproducible research including data sharing [32].

In the attempt of reproducing analyses from RCTs, Naudet et. al experienced that researchers requested financial compensation for preparing data for sharing [35]. Evidence from data access inquiries in individual patient data for meta-analysis suggests that financial incentives, however, is not effective [46].

In our view, a more valuable approach is to share data in a repository, making it accessible without the need for authors to reply to messages, and incorporating data preparation as part of the publication process. However, this approach remains underrepresented [34, 37]. Imposing a requirement to deposit data and models in a repository prior to the publication of a manuscript will help against data loss and will render the argument about lack of resources for data sharing invalid. To be fully effective this would need to be checked by the publishers. Such a requirement will increase the work load for the authors as well as for the publisher, but the increased work load will be mainly limited to the time around the submission.

With access restrictions bound to a single author, though, this will still not help against email decay or unresponsiveness. Therefore, we argue an automatic maximal embargo time needs to be enforced after which the data will become openly accessible. Data accessibility should be especially of interest for funders who payed for the data generation and model development. These are often public entities or trusts that should have an incentive to make the most of their investment. The funders also have the means to put enough pressure upon the involved parties to adhere to data and model publications rules by excluding researchers who do not follow these rules from further grant applications (as is done [47]). The role of the funders with respect to data sharing has been evaluated with the result that funders ought to promote data sharing and that making data sharing mandatory is justifyable in certain situations [48]. First steps in this direction have been taken. For instance, in 2015 the National Institutes of Health (NIH) published a public plan to promote access to scientific publications which states: “NIH intends to make public access to digital scientific data the standard for all NIH-funded research” [49]. We did not see this stated intention being acted upon in our case study, as access to research data from NIH-funded projects was not granted in our case example. Still, we believe that promoting data sharing through funders is the route with the highest probabilty for success in increasing the levels of data sharing.

## 5 Conclusion

It is well established that making data and trained models available is crucial for scientific progress. Moreover, making data and trained models available enables the validation and adaptation of models for specific clinical applications, ultimately facilitating their integration into clinical routine care. Despite the long-standing demands for reproducible research from the scientific community and efforts to incentivize data sharing, our case study reveals a persistent lack of data sharing culture. This is particularly concerning given the potential adaptability of DL models. The reasons for this lack of data sharing are multifaceted and include, but are not limited to, limited resources, privacy constraints, and unresponsiveness of authors. To overcome these barriers, we propose alternative mechanisms to foster a data sharing culture that rewards researchers that conduct reproducible research.

## List of abbreviations

AI: Artificial Intelligence AUC: Area Under the Curve
DAS: Data availability statement DL: Deep learning
ML: Machine learning
NIH: National Institutes of Health LLM: Large Language Model

## Supplementary information

- Supplement 1: PubMed Search Query
- Supplement 2: Study inclusion process flow chart
- Supplement 3: Detailed results in the form of an excel spreadsheet (Study selection process, Characteristics of the included studies, Data inquiry)
- Supplement 4: R Source code for analysis and creation of graphics

## Declarations

### Ethics approval and consent to participate

Not applicable. Clinical trial number: not applicable.

### Consent for publication

Not applicable. This study does not contain any individual person’s data that is not publicly available. The data presented in this study is either publicly available through the published information alongside the included studies (author names and email addresses) or has been anonymized to protect individual identities. Any responses to our inquiries are presented in a summarised version.

### Availability of data and materials

All data generated or analysed during this study are included in this published article and its supplementary information files.

## Author contribution

MS screened and reviewed the literature, extracted the data from publications, performed the data inquiry process and the statistical analysis. AL supported the data inquiry process. Both MS and AL contributed equally in writing the manuscript. All authors read and approved the final manuscript.

## Supporting information

Supplement 1: PubMed Search Query

Supplement 2: Study inclusion process flow chart

Supplement 3: Detailed results in the form of an excel spreadsheet (Study selection process, Characteristics of the included studies, Data inquiry)

Supplement 4: R Source code for analysis and creation of graphics

## Data Availability

All data produced in the present work are contained in the manuscript

## Acknowledgements

We thank Judith Heinz for the support in the preparation of the PubMed Search Query, Josef Hammoud for the support in cross-checking the results of the data extraction and Wendi Qu for the quality control of the supplementary material.

## Declaration of generative AI and AI-assisted technologies in the writing process

During the preparation of this work the author(s) used a locally-hosted version of LLaMA, in both variants 8B and 70B, for proofreading and text optimization of the manuscript, and support in the development of source code for creating tables and plots. After using this tool/service, the author(s) reviewed and edited the content as needed and take(s) full responsibility for the content of the published article.

## Appendix A Details on the literature search

### A.1 Search strategy

The search strategy was designed to identify PubMed articles published in the last 10 years, in English, combining three main concepts: artificial intelligence methods, cervical carcinoma, and colposcopy images. We restricted our search to the last 10 years to focus on recent advances in AI-based analyses, and to English language publications to ensure clarity and consistency in the identified studies. The artificial intelligence methods component included terms such as ”deep learning”, ”machine learning”, and ”artificial intelligence”, searched in both the title and abstract, as well as in the MeSH terms. The cervical carcinoma component included terms such as ”cervical carcinoma”, ”cervix carcinoma”, and combinations of words like ”cervical” and ”carcinoma”, searched in the title and abstract. The colposcopy images component included terms like ”colposcopy” in MeSH terms and title/abstract, and combinations of words like ”colposcopic” and ”image”, searched in the title and abstract. The three components were combined using the AND operator to retrieve articles that discuss the diagnosis of cervical carcinoma using artificial intelligence methods and involve colposcopy images. The date of the last search was 5 May 2024. The full search query is provided in Supplement 1.

### A.2 Study selection

The list of abstracts was screened for inclusion and full-text versions of all articles identified as potentially relevant were evaluated by one reviewer (MS) using the following inclusion criteria: (i) an application of ML methods, (ii) using images from colposcopy, and (iii) with the intent to support treatment decisions for cervical carcinoma. We included studies that developed or validated AI-based models for cervical carcinoma classification or segmentation. We excluded studies that provided systematic reviews and/or overviews of existing literature in the field, as they did not contribute to the development or validation of AI-based models for cervical carcinoma diagnosis. Further, we excluded studies that met any of the following criteria: (i) where the ground truth for colposcopy image was based on expert opinion, (ii) the classification task was not relevant to CIN classification of cervical carcinoma (e.g., quality classification of the cervical image), or (iii) the study had too few data (fewer than 200 images), as this sample size was considered too small to provide reliable results. In addition to the PubMed Query, we identified studies by citation that were either used as data sources in one or multiple of the primary studies or that explicitly provide a dataset of colposcopy images, and we included these studies in this work.

### A.3 Data extraction and analysis

One reviewer (MS) extracted additional information related to the included studies. We categorized each publication as either an analysis or publication that provides data, i.e. was referenced by one of the primary studies as data source. For analysis publications, we further identified the task performed (segmentation or classification) and, for classification tasks, the specific classification type (binary or multiclass or both). We also gathered information on the data used, including the source of the data (specific external data source or otherwise), country of data collection, dataset size, and class distribution of images in the dataset (computed as the number of cases on the tital number of images in the dataset, for binary classification studies only). For classification tasks, we retrieved the performance metrics Area Under the Receiver Operating Characteristic Curve (AUC) (C-statistic) and Accuracy. When multiple models were compared, we recorded the performance metric of the best-performing model, considering only the model’s performance (excluding expert opinion or other factors) and only on test or validation datasets.

## Appendix B Further characteristics of the included studies

### B.1 Performed Tasks

The analysis studies focused on various tasks, with the majority (27) performing classification tasks, 6 performing segmentation tasks, and 6 undertaking both. The classification tasks aimed to classify stages of cervical cancer, while segmentation tasks involved the automated identification and demarcation of specific regions or features within the colposcopy images, ultimately assisting in the diagnosis and grading of cervical cancer.

Among the 33 studies that performed classification tasks, 21 used binary classification, 6 employed multiclass classification, and 6 attempted both. These studies utilized either the World Health Organization (WHO) grading system, which distinguishes between low-grade squamous intraepithelial lesions (LSIL) and high-grade squamous intraepithelial lesions (HSIL) for classification tasks (16/33), or the cervical intraepithelial neoplasia (CIN) classification system (14/33). For three studies, the class distinction methodology was unclear.

Among the 27 studies that incorporated binary classification tasks, 5 studies performed two different classifications, resulting in a total of 32 attempts to classify binary outcomes. The binary classification tasks aimed at identifying patients with HSIL or more severe (HSIL+/CIN2+) in 18 cases, and patients with LSIL or more severe (LSIL+/CIN1+) in 5 cases. Beyond that, the studies attempted to classify the CIN3+ cases (2), determine the Need-to-Biopsy (1), identify Precancer+ cases (2), differentiate chronic cervicitis from cancer (1), or simply distinguish between LSIL and HSIL (3).

Among the 12 studies that employed multiclassification, a range of classification systems were utilized. The most common system, used by 4 studies, classified images into 4 categories: Normal, CIN1, CIN2, CIN3, and Cancer, of which 3 combined CIN2/3 to one class. One study omitted the Cancer category, instead distinguishing only between Normal, CIN1, CIN2, and CIN3. Besides the CIN-classification system, studies used WHO-classes Normal, LSIL, HSIL as a basis for classification (4), of which one further classified HSIL+, one further included Cancer (1) and Neoplasms (1) as categories. Other multiclassifications involved Normal, gray zone, precancer+ (2), and severe dysplasia, carcinoma in situ (CIS) and invasive cancer (IC) (1).

### B.2 External data sources used

**Table B1.** Overview on external data sources used for training AI models.

| Study | Description | Origin of the data | Number of images |
| --- | --- | --- | --- |
| Guanacaste project[1] | Also referred to as the Natural History Study (NHS), a population-based prospective study conducted in Guanacaste, Costa Rica, between 1993 and 2000. The study aimed to investigate the role of human papillomavirus (HPV) infection and its co-factors in the etiology of high-grade cervical neoplasia, and to evaluate new cervical cancer screening technologies. The dataset contains information on a cohort of women who underwent screening visits, including collection of specimens for cytology, HPV testing, and images of the cervix, as well as histology among women with abnormal colposcopic evaluation. | Costa Rica | 9,901 |
| ALTS[2] | Also referred to as ASCUS/LSIL Triage Study for Cervical Cancer (ALTS), a multi-center randomized trial conducted between 1996 and 2000 with the aim to compare strategies in handling of low-grade squamous intraepithelial lesion (LSIL) cytology. This study enrolled US women with a LSIL interpretation and randomly assigned participants into three different management strategies: immediate colposcopy, triage based on HPV DNA testing and liquid-based cytology or repeated cytology. Women were followed for 2 years with screening visits every 6 months. | USA | 4,027 |
| Costa Rica Vaccine Trial (CVT)[3] | The CVT study was a double-blind, randomized-controlled trial investigating the efficacy of human papillomavirus vaccine compared to hepatitis A vaccine in the prevention of cervical intraepithelial neoplasia or cervical cancer in healthy young adult women in Costa Rica, Guanacaste, and parts of the Puntarenas provinces. | Costa Rica | 1,176 |
| Kaggle Dataset [4] | This dataset is publicly available through the Kaggle platform as part of the "Intel & MobileODT Cervical Cancer Screening Competition". It comprises images of the cervix captured using a smartphone-based digital cervical imaging device. The primary goal of the challenge was to develop a classification algorithm that can categorize these images into the three distinct cervix types, identifying the location of the transformation zone. This enables the identification of women who require further testing for cervical cancer. | unclear | 8,727 |
| Biopsy study (Biop)[5] | The Biopsy study refers to a population-based study of women referred to colposcopy for abnormal cervical cancer screening results at the University of Oklahoma Health Sciences Center from 2009 to 2011. The study aimed to investigate the accuracy of a combined p16 and Ki-67 cytology for detection of cervical precancer. All women enrolled had a colposcopy and at least one biopsy. | USA | 737 |
| Biopsy Study - Europe (D Biop)[6] | The European Biopsy study refers to a study of women referred for colposcopy and diagnosed with CIN2/3 or invasive cervical cancer, investigating high-risk HPV genotypes in these women. | Spain, Netherlands | 1,172 |
| DYSIS[7] | The DYSIS study was a prospective, comparative, multi-centre clinical trial to validate the dynamic spectral imaging (DSI) colposcope's color-coded map in discriminating high- from low-grade cervical lesions and non-neoplastic tissue in women referred for colposcopy. Images of the cervix were captured during routine colposcopy examinations using the DYSIS digital colposcope. The images are accompanied by biopsy results. | Netherlands | 1,100 |

**Table B2.** Reported performance metrics AUC (C-statistic) and Accuracy for studies performing binary and multiclassification tasks.

|  | Min | 25% | 50%, | 75% | Max | Mean |
| --- | --- | --- | --- | --- | --- | --- |
| Binary AUC | 0.674 | 0.765 | 0.857 | 0.91 | 0.98 | 0.838 |
| Binary Accuracy | 0.630 | 0.807 | 0.823 | 0.886 | 0.9519 | 0.8297 |
| Multiclass AUC | 0.777 | 0.965 | 0.97 | 0.9856 | 0.99 | 0.9375 |
| Multiclass Accuracy | 0.435 | 0.4965 | 0.6669 | 0.9156 | 0.9687 | 0.6953 |

### B.3 Details on the data used

For those studies with their own datasets and the publications providing data (excluding the publicly available dataset), in total 39 studies, the majority of the data was collected in China (in 19 cases), followed by Korea (5) and Japan (4). Therefore, the majority of data comes from an Asian population (with India, 29 studies). This is only followed by four studies with data origin in North America (USA and Costa Rica), two studies from Europe (Netherlands, Spain), and two studies with data origin in Africa (Zambia, Nigeria). Two studies reported multiple data origin countries and were therefore classified as “Various”.

The number of colposcopy images used in the 39 analysis studies varied widely (Minimum 213; Maximum 101,267). Notably, studies that validated pre-trained models used smaller numbers of images, with 5 publications performing validation (Minimum 213; Maximum 7,457). In contrast, studies that trained their own models used a wide range of images, from 253 to 101,267 in total. It is important to note that multiple images may exist for a single patient, captured at different time slots after acetic acid application, or with different filters or quality. Half of the studies that trained their own models relied on a dataset of between 1,813 and 12,990 images.

However, the amount of data is only one aspect of developing reliable models. The class distribution, particularly in classification tasks, is equally crucial. In the case of binary classification, the proportion of cases (such as LSIL+ or HSIL+) on the total number of images varied notably, ranging from 3.77% to 83%. For half of the studies, this proportion fell between 17.48% (1st Quartil) and 52.6% (3rd Quartil). This imbalance in data for some of the included studies may have a profound impact on the performance of these models, potentially leading to biased or inaccurate results.

### B.4 Reported performance measures

Among the 27 studies performing binary classification tasks, 8 out of 41 did not report C-statistics and 11 did not report on accuracy measures. Similarly, for those performing multiclassification tasks, 7 out of 12 did not report on C-statistics and 3 did not report on accuracy. However, for the studies that did provide performance measures, some demonstrated exceptional discrimination performance. Table B2 provides a summary of the reported AUC and Accuracy for studies performing classification tasks. If a study performed separate classification tasks, each reported performance metrics is included. Specifically, the top 25% of binary classification studies achieved an AUC of 0.91 or higher, while the top 25% of multiclassification studies reached an AUC of 0.986 or higher. Accuracy is reported similarly for binary classifications, and, in general, tends to be lower for multiclassifications.

## Appendix C List of included studies

