## Supplement 1: PubMed Search Query for "A Case Study on Colposcopy-Based Cervical Cancer Staging Reveals an Alarming Lack of Data Sharing Hindering the Adoption of Machine Learning in Clinical Practice"

**Classification of cervical carcinoma based on colposcopy images using artificial intelligence methods**

**Search Strategy – PubMed (Suche 03.05.2024)**

| SEARCH | QUERY |
| --- | --- |
| #1 | “deep learning”[Title/Abstract] OR deep learning[MESH Terms] |
| #2 | “machine learning”[Title/Abstract] OR machine learning[MESH Terms] |
| #3 | “artificial intelligence”[Title/Abstract] OR artificial intelligence[MESH Terms] |
| #4 | “hierarchical learning”[Title/Abstract] |
| #5 | “decision support models”[Title/Abstract] OR “decision support model”[Title/Abstract] |
| #6 | “machine intelligence”[Title/Abstract] |
| #7 | #1 OR #2 OR #3 OR #4 OR #5 OR #6 |
| #8 | uterine cervical neoplasms[MESH Terms] |
| #9 | “cervical carcinoma”[Title/Abstract] OR (cervical[Title/Abstract] AND (carcinoma[Title/Abstract] OR cancer[Title/Abstract] OR neoplasm[Title/Abstract] OR neoplasms[Title/Abstract] OR neoplasia[Title/Abstract] OR dysplasia[Title/Abstract] OR lesion[Title/Abstract] OR lesions[Title/Abstract] OR tumor[Title/Abstract] OR tumors[Title/Abstract] OR tumour[Title/Abstract] OR tumours[Title/Abstract])) |
| #10 | “cervix carcinoma”[Title/Abstract] OR (cervix[Title/Abstract] AND (carcinoma[Title/Abstract] OR cancer[Title/Abstract] OR neoplasm[Title/Abstract] OR neoplasms[Title/Abstract] OR neoplasia[Title/Abstract] OR dysplasia[Title/Abstract] OR lesion[Title/Abstract] OR lesions[Title/Abstract] OR tumor[Title/Abstract] OR tumors[Title/Abstract] OR tumour[Title/Abstract] OR tumours[Title/Abstract])) |
| #11 | #9 OR #10 |
| #12 | (cervical[Title/Abstract] AND screening[Title/Abstract]) OR (cervix[Title/Abstract] AND screening[Title/Abstract]) |
| #13 | colposcopy[MESH Terms] |
| #14 | colposcopy[Title/Abstract] OR colposcopies[Title/Abstract] OR (colposcopic[Title/abstract] AND (image[Title/Abstract] OR images[Title/Abstract])) |
| #15 | #12 OR #13 OR #14 |
| #16 | #7 AND #11 AND #15 |
| #17 | #16 Filters: published in the last 10 years; English |
