## Supplementary figures and images for "A Case Study on Colposcopy-Based Cervical Cancer Staging Reveals an Alarming Lack of Data Sharing Hindering the Adoption of Machine Learning in Clinical Practice"

### Supplement 2: Study inclusion process flow chart

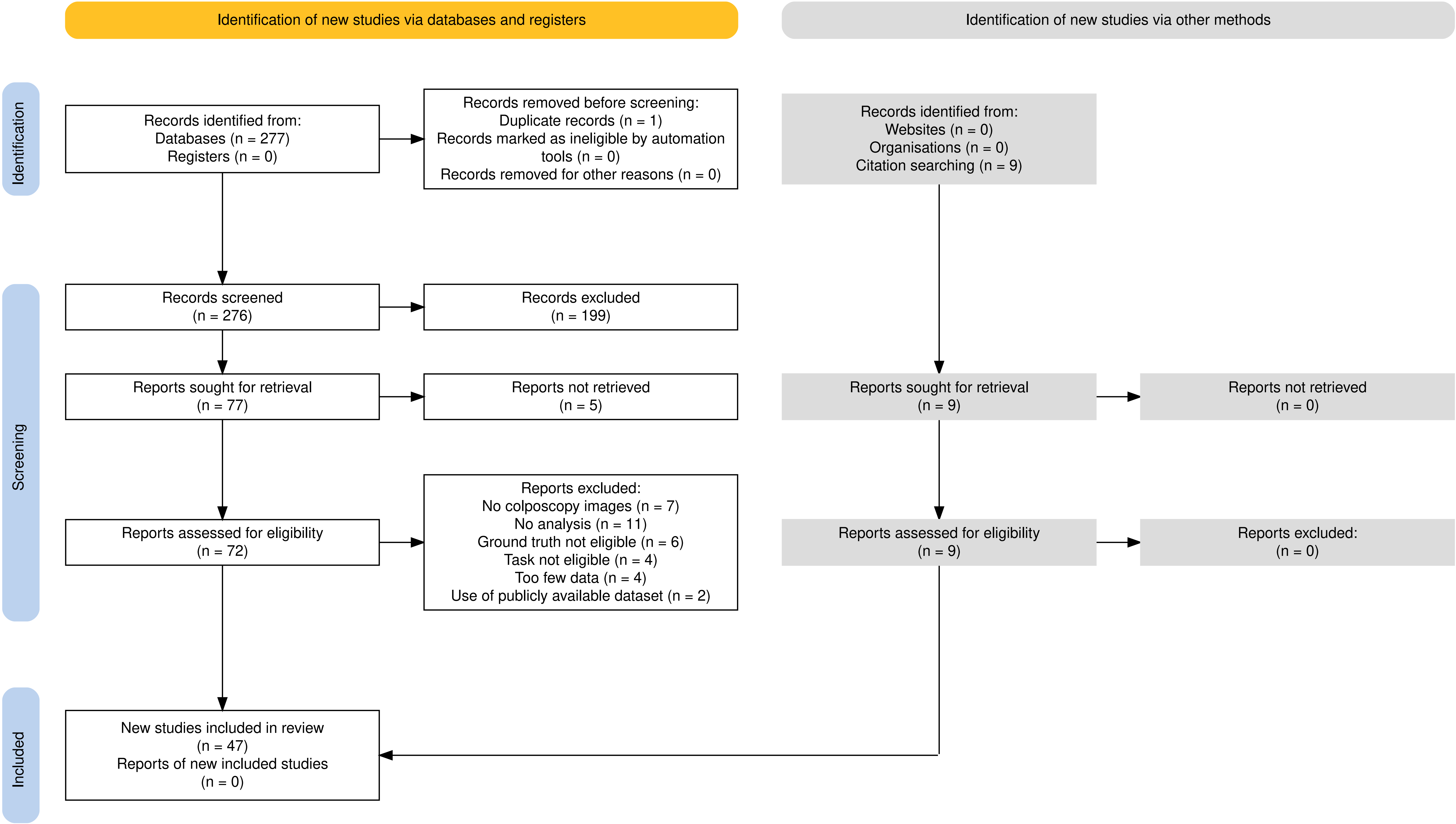
