## Supplement 4: R Source code for analysis and creation of graphics for "A Case Study on Colposcopy-Based Cervical Cancer Staging Reveals an Alarming Lack of Data Sharing Hindering the Adoption of Machine Learning in Clinical Practice": 3.2_Characteristics and data availability of included studies.R

```
# setwd() # set appropriate working directory

library(readxl)
library(dplyr)
library(tidyverse)

data <- read_xlsx("Supplement3_Detailed_results.xlsx", sheet = "analysis_table" )

### Type of publication
# -------------------------------------

type_of_publication <- data %>%
  group_by(`Type of publication`) %>%
  summarise(Count = n()) 

paste("Number of analysis publications:", type_of_publication$Count[type_of_publication$`Type of publication` == "Analysis publication"])
paste("Number of publications providing data:", type_of_publication$Count[type_of_publication$`Type of publication` == "Publication providing data"])
paste("Number of publicly available datasets:", type_of_publication$Count[type_of_publication$`Type of publication` == "Publicly available dataset"])

### Use of external data sources
# --------------------------------------

use_external_data <- data %>% 
  filter(`Type of publication` == "Analysis publication") %>% 
  group_by(`Use of external data sources (Y/N)`) %>%
  summarise(Count = n())

paste("Number of studies using external data sources:", use_external_data$Count[use_external_data$`Use of external data sources (Y/N)` == "Y"])

external_data_count <- data %>%
  filter(`Use of external data sources (Y/N)` == "Y") %>%
  pivot_longer(cols = c(`Guanacaste project (Y/N)`, `Costa Rica Vaccine Trial (CVT) (Y/N)`, `ALTS (Y/N)`, `Kaggle Dataset (Y/N)`, `Biopsy Study - Europe (D Biop) (Y/N)`,
                        `Biopsy study (Biop) (Y/N)`, `DYSIS (Y/N)`),
               names_to = "Dataset",
               values_to = "Used") %>%
  filter(Used == "Y") %>%
  mutate(Dataset = gsub(" \\(Y/N\\)", "", Dataset)) %>%
  group_by(Dataset) %>%
  summarise(Count = n()) 

paste("Number of studies using data from the Guanacaste project:", external_data_count$Count[external_data_count$Dataset == "Guanacaste project"])
paste("Number of studies using data from the ALTS study:", external_data_count$Count[external_data_count$Dataset == "ALTS"])
paste("Number of studies using data from the Costa Rica Vaccine Trial:", external_data_count$Count[external_data_count$Dataset == "Costa Rica Vaccine Trial (CVT)"])
paste("Number of studies using data from the Kaggle dataset:", external_data_count$Count[external_data_count$Dataset == "Kaggle Dataset"])
paste("Number of studies using data from the Biopsy study:", external_data_count$Count[external_data_count$Dataset == "Biopsy study (Biop)"])
paste("Number of studies using data from the Biopsy study - Europe:", external_data_count$Count[external_data_count$Dataset == "Biopsy Study - Europe (D Biop)"])
paste("Number of studies using data from the DYSIS:", external_data_count$Count[external_data_count$Dataset == "DYSIS"])

paste("Number of studies not using external data sources:", use_external_data$Count[use_external_data$`Use of external data sources (Y/N)` == "N"])

### Data availability statement 
# ---------------------------------------

das_available <- data %>% filter(`Type of publication` != "Publicly available dataset") %>%
  group_by(`DAS available (Y/N)`) %>%
  summarise(Count = n())

paste("Number of studies with DAS:", das_available$Count[das_available$`DAS available (Y/N)` == "Y"])
paste("Number of studies without DAS:", das_available$Count[das_available$`DAS available (Y/N)` == "N"])

# DAS for external data sources
data %>% filter(`Type of publication` == "Publication providing data") %>%
  group_by(`DAS available (Y/N)`) %>%
  summarise(Count = n())

# studies with DAS: own or external data sources
das_available_dataset <- data %>% filter(`DAS available (Y/N)` == "Y") %>%
  group_by(`Use of external data sources (Y/N)`) %>%
  summarise(Count = n())

paste("Number of studies with DAS that use own dataset:", das_available_dataset$Count[das_available_dataset$`Use of external data sources (Y/N)` == "N"])
paste("Number of studies with DAS that use external data sources:", das_available_dataset$Count[das_available_dataset$`Use of external data sources (Y/N)` == "Y"])

### Data availability statement over time
# ---------------------------------------

DAS_over_time <- data %>%
  filter(`Type of publication` != "Publicly available dataset") %>%
  mutate(Year = ifelse(Year < 2018, "<2018", as.character(Year))) %>%
  group_by(Year) %>%
  summarise(
    Studies_including_DAS = sum(`DAS available (Y/N)` == "Y"),
    Total_number_studies = n()) %>%
  mutate(Proportion = (Studies_including_DAS / Total_number_studies)*100)
print(DAS_over_time)

### Generic data availability statement
# ----------------------------------------

generic_das <- data %>% filter(`Type of publication` != "Publicly available dataset",
                               `DAS available (Y/N)` == "Y") %>%
  group_by(`Generic DAS with data available on request`) %>%
  summarise(Count = n())

paste("Number of studies with generic DAS:", generic_das$Count[generic_das$`Generic DAS with data available on request` == "Y"])
paste("Number of studies with specific DAS:", generic_das$Count[generic_das$`Generic DAS with data available on request` == "N"])

### Raw data availability 
# ----------------------------------------

DAS_rawdata <- data %>% filter(`Type of publication` != "Publicly available dataset",
                               `DAS available (Y/N)` == "Y", 
                               `Generic DAS with data available on request` == "N") %>%
  group_by(`Raw data available (Y/N)`) %>%
  summarise(Count = n()) 

new_row <- data.frame(`Raw data available (Y/N)` = "Not mentioned", Count = 0)
colnames(new_row) <- c("Raw data available (Y/N)", "Count")

DAS_rawdata <-  DAS_rawdata %>%
  rbind(new_row)

paste("Number of studies declaring raw data available:", DAS_rawdata$Count[DAS_rawdata$`Raw data available (Y/N)` == "Y"])
paste("Number of studies declaring raw data unavailable:", DAS_rawdata$Count[DAS_rawdata$`Raw data available (Y/N)` == "N"])
paste("Number of studies not mentioning the availability of raw data:", DAS_rawdata$Count[DAS_rawdata$`Raw data available (Y/N)` == "Not mentioned"])

DAS_rawdata_accessibility <- data %>% filter(`Type of publication` != "Publicly available dataset",
                `DAS available (Y/N)` == "Y", 
                `Generic DAS with data available on request` == "N",
                `Raw data available (Y/N)` == "Y") %>%
  group_by(`Accessibility of raw data`) %>%
  summarise(Count = n())

paste("Number of studies declaring raw data available upon request:", DAS_rawdata_accessibility$Count[DAS_rawdata_accessibility$`Accessibility of raw data` == "On request"])

### Source code availability 
# ----------------------------------------

DAS_code <- data %>% filter(`Type of publication` != "Publicly available dataset",
                            `DAS available (Y/N)` == "Y", 
                            `Generic DAS with data available on request` == "N") %>%
  group_by(`Code available (Y/N)`) %>%
  summarise(Count = n()) 

new_row <- data.frame(`Code available (Y/N)` = "N", Count = 0)
colnames(new_row) <- c("Code available (Y/N)", "Count")

DAS_code <- DAS_code %>%
  rbind(new_row)

paste("Number of studies declaring source code available:", DAS_code$Count[DAS_code$`Code available (Y/N)` == "Y"])
paste("Number of studies declaring source code unavailable:", DAS_code$Count[DAS_code$`Code available (Y/N)` == "N"])
paste("Number of studies not mentioning the availability of source code:", DAS_code$Count[DAS_code$`Code available (Y/N)` == "Not mentioned"])

#accessibility of source code
DAS_code_accessibility <- data %>% filter(`Type of publication` != "Publicly available dataset",
                                             `DAS available (Y/N)` == "Y", 
                                             `Generic DAS with data available on request` == "N",
                                          `Code available (Y/N)` == "Y") %>%
  group_by(`Accessibility of code`) %>%
  summarise(Count = n())

paste("Number of studies declaring source code available with link:", DAS_code_accessibility$Count[DAS_code_accessibility$`Accessibility of code` == "Directly available"])

### Pre-trained weights availability
# ------------------------------------------

DAS_weights <- data %>% filter(`Type of publication` != "Publicly available dataset",
                               `DAS available (Y/N)` == "Y", 
                               `Generic DAS with data available on request` == "N") %>%
  group_by(`Pre-trained weights available (Y/N)`) %>%
  summarise(Count = n()) 

new_row <- data.frame(`Pre-trained weights available (Y/N)` = c("Y", "N"), Count = c(0,0))
colnames(new_row) <- c("Pre-trained weights available (Y/N)", "Count")

DAS_weights <- DAS_weights %>%
  rbind(new_row)

paste("Number of studies declaring pre-trained weights available:", DAS_weights$Count[DAS_weights$`Pre-trained weights available (Y/N)` == "Y"])
paste("Number of studies declaring pre-trained weights unavailable:", DAS_weights$Count[DAS_weights$`Pre-trained weights available (Y/N)` == "N"])
paste("Number of studies not mentioning the availability of pre-traiend weights:", DAS_weights$Count[DAS_weights$`Pre-trained weights available (Y/N)` == "Not mentioned"])

### Positiv/negative DAS
# --------------------------------------------

DAS_positiv_negativ <- data %>% filter(`Type of publication` != "Publicly available dataset",
                `DAS available (Y/N)` == "Y") %>%
  group_by(`Data declared available (Y/N)`) %>%
  summarise(Count = n())

paste("Number of studies with positive DAS:", DAS_positiv_negativ$Count[DAS_positiv_negativ$`Data declared available (Y/N)` == "Y"])
paste("Number of studies with negative DAS:", DAS_positiv_negativ$Count[DAS_positiv_negativ$`Data declared available (Y/N)` == "N"])

DAS_positiv_generic <- data %>% filter(`Type of publication` != "Publicly available dataset",
                                       `DAS available (Y/N)` == "Y",
                                       `Data declared available (Y/N)` == "Y") %>%
  group_by(`Generic DAS with data available on request`) %>%
  summarise(Count = n())

paste("Number of studies with positive generic DAS:", DAS_positiv_generic$Count[DAS_positiv_generic$`Generic DAS with data available on request` == "Y"])
paste("Number of studies with positive specific DAS:", DAS_positiv_generic$Count[DAS_positiv_generic$`Generic DAS with data available on request` == "N"])
```
