## Supplement 4: R Source code for analysis and creation of graphics for "A Case Study on Colposcopy-Based Cervical Cancer Staging Reveals an Alarming Lack of Data Sharing Hindering the Adoption of Machine Learning in Clinical Practice": 3.3_Data sharing outcomes and responses.R

```
# setwd() # set appropriate working directory

library(readxl)
library(dplyr)
library(tidyverse)

data <- read_xlsx("Supplement3_Detailed_results.xlsx", sheet = "Data inquiry details")
analysis_data <- read_xlsx("Supplement3_Detailed_results.xlsx", sheet = "analysis_table")


### Subject of inquiry
# -------------------------------------

subject_inquiry <- analysis_data %>% 
  filter(`Type of publication` != "Publicly available dataset") %>%
  group_by(`Subject of inquiry`) %>%
  summarise(Count = n())

paste("Number of studies requested for access to dataset:", subject_inquiry$Count[subject_inquiry$`Subject of inquiry` == "Dataset"])
paste("Number of studies requested for access to dataset + pre-trained model:", subject_inquiry$Count[subject_inquiry$`Subject of inquiry` == "Dataset + pre-trained model"])
paste("Number of studies requested for access to pre-trained model:", subject_inquiry$Count[subject_inquiry$`Subject of inquiry` == "Pre-trained model"])

### Initial Email inquiry 
# -------------------------------------

# Number of studies that we inquired via mail (first inquiry)
studies_with_initial_mail_inquiry <- data %>%
  group_by(`Pub ID`) %>%
  summarise(has_mail_inquiry = any(`Date of 1st inquiry via mail` != "N/A"))

paste("Number of studies contacted via initial email inquiry:", nrow(studies_with_initial_mail_inquiry[studies_with_initial_mail_inquiry$has_mail_inquiry == T,]))

# Count number of authors contacted via mail by type
authors_contacted_via_mail <- data %>%
  filter(`Date of 1st inquiry via mail` != "N/A") %>%
  group_by(`Pub ID`, `Type of author`) %>%
  summarise(count = n(), .groups = "drop") %>%
  pivot_wider(
    names_from = `Type of author`,
    values_from = count,
    values_fill = 0,
    id_cols = `Pub ID`
  ) %>%
  mutate(
    total_authors = `Corresponding author + First author` + `Other` + `Corresponding author` + `First author`
  )

# Calculate total counts for each author type
corresponding_author_count <- sum(authors_contacted_via_mail$`Corresponding author`)
corresponding_author_first_count <- sum(authors_contacted_via_mail$`Corresponding author + First author`)
first_author_count <- sum(authors_contacted_via_mail$`First author`)
other_author_count <- sum(authors_contacted_via_mail$Other)
total_authors_count <- sum(authors_contacted_via_mail$total_authors)

paste("Corresponding author count:", corresponding_author_count)
paste("Corresponding author + First author count:", corresponding_author_first_count)
paste("First author count:", first_author_count)
paste("Other author count:", other_author_count)
paste("Total authors count:", total_authors_count)

# Count unique authors contacted
unique_authors_contacted <- data %>%
  filter(`Pub ID` != "45") %>%
  distinct(`Name of author`, .keep_all = TRUE) %>%
  group_by(has_mail_inquiry = `Date of 1st inquiry via mail` != "N/A") %>%
  summarise(Count = n()) %>%
  filter(has_mail_inquiry)

paste("Number of unique authors contacted:",unique_authors_contacted$Count)

# Responses to first inquiry via mail
responses_first_inquiry <- data %>%
  filter(`Date of 1st inquiry via mail` != "N/A") %>%
  mutate(response_first = case_when(
    `Response to 1st inquiry via mail` == "Mail address not available anymore" ~ "Failed",
    `Response to 1st inquiry via mail` == "No response" ~ "No response",
    TRUE ~ "Response"
  )) %>%
  group_by(`Pub ID`, response_first) %>%
  summarise(count = n()) %>%
  pivot_wider(
    values_from = count,
    names_from = response_first,
    id_cols = `Pub ID`,
    values_fill = 0
  ) %>%
  mutate(
    response_per_study = case_when(
      Response >= 1 ~ "Response",
      Failed >= 1 & Response == 0 & `No response` == 0 ~ "Failed",
      TRUE ~ "No response"
    )
  )

# Summarize response counts
failed_count <- sum(responses_first_inquiry$Failed)
response_count <- sum(responses_first_inquiry$Response)
no_response_count <- sum(responses_first_inquiry$`No response`)

paste("Failed count:", failed_count)
paste("Response count:", response_count)
paste("No response count:", no_response_count)

# Responses to first inquiry via mail on unique authors level
unique_author_responses <- data %>%
  filter(`Pub ID` != "45", `Date of 1st inquiry via mail` != "N/A") %>%
  distinct(`Name of author`, .keep_all = TRUE) %>%
  group_by(`Response to 1st inquiry via mail`) %>%
  summarise(Count = n())

failed_count_unique <- unique_author_responses$Count[unique_author_responses$`Response to 1st inquiry via mail` == "Mail address not available anymore"] 
response_count_unique <- unique_author_responses$Count[unique_author_responses$`Response to 1st inquiry via mail` == "Pending response"] +
  unique_author_responses$Count[unique_author_responses$`Response to 1st inquiry via mail` == "Redirect/Referral response"] + 
  unique_author_responses$Count[unique_author_responses$`Response to 1st inquiry via mail` == "Data sharing declined"] 
no_response_count_unique <- unique_author_responses$Count[unique_author_responses$`Response to 1st inquiry via mail` == "No response"]  

paste("Failed count (unique authors):", failed_count_unique)
paste("Response count (unique authors):", response_count_unique)
paste("No response count (unique authors):", no_response_count_unique)

# Responses on study-level
study_response_counts <- responses_first_inquiry %>%
  group_by(response_per_study) %>%
  summarise(count = n())

paste("Failed count (study-level):", study_response_counts$count[study_response_counts$response_per_study == "Failed"])
paste("Response count (study-level):", study_response_counts$count[study_response_counts$response_per_study == "Response"])
paste("No response count (study-level):", study_response_counts$count[study_response_counts$response_per_study == "No response"])

### Follow-up Email inquiry
# -------------------------------------

# Number of studies with follow-up email inquiry
studies_with_followup_mail <- data %>%
  # for those studies that had not answered after first inquiry:
  filter(`Pub ID` %in% as.vector(t(responses_first_inquiry %>% filter(response_per_study == "No response") %>% select(`Pub ID`)))) %>% 
  group_by(`Pub ID`) %>%
  summarise(has_followup_mail = any(`Date of 2nd inquiry via mail`!= "N/A"))

paste("Number of studies followed up via mail:", nrow(studies_with_followup_mail[studies_with_followup_mail$has_followup_mail == T,]))

# Number of contacted authors
second_inquiry_contacts <- data %>%
  # for those studies that had not answered after first inquiry:
  filter(`Pub ID` %in% as.vector(t(responses_first_inquiry %>% filter(response_per_study == "No response") %>% select(`Pub ID`)))) %>% 
  filter(`Date of 2nd inquiry via mail` != "N/A") %>%
  group_by(`Pub ID`, `Type of author`) %>%
  summarise(count = n(), .groups = "drop") %>%
  pivot_wider(
    names_from = `Type of author`,
    values_from = count,
    values_fill = 0,
    id_cols = `Pub ID`
  ) %>%
  mutate(
    total_authors = `Corresponding author + First author` + `Other` + `Corresponding author` + `First author`
  )

# Summarize contact counts
total_contacts_count <- sum(second_inquiry_contacts$total_authors)
corresponding_author_count <- sum(second_inquiry_contacts$`Corresponding author`)
corresponding_author_first_count <- sum(second_inquiry_contacts$`Corresponding author + First author`)
first_author_count <- sum(second_inquiry_contacts$`First author`)
other_author_count <- sum(second_inquiry_contacts$`Other`)

paste("Total contacts count:", total_contacts_count)
paste("Corresponding author count:", corresponding_author_count)
paste("Corresponding author + First author count:", corresponding_author_first_count)
paste("First author count:", first_author_count)
paste("Other author count:", other_author_count)

# Count unique authors contacted
unique_authors_contacted_second <- data %>%
  filter(`Pub ID` != "45") %>%
  distinct(`Name of author`, .keep_all = TRUE) %>%
  group_by(has_followup_mail = `Date of 2nd inquiry via mail` != "N/A") %>%
  summarise(Count = n()) %>%
  filter(has_followup_mail)

paste("Number of unique contacted authors via follow up email:", unique_authors_contacted_second$Count)

# Responses to follow up mail
second_inquiry_responses <- data %>%
  filter(`Date of 1st inquiry via mail` != "N/A") %>%
  mutate(
    response_first = case_when(
      `Response to 1st inquiry via mail` == "Mail address not available anymore" ~ "Failed",
      `Response to 1st inquiry via mail` == "No response" ~ "No response",
      TRUE ~ "Response"
    ),
    response_second = case_when(
      `Response to 2nd inquiry via mail` == "Mail address not available anymore" ~ "Failed",
      `Response to 2nd inquiry via mail` == "No response" ~ "No response",
      TRUE ~ "Response"
    )
  ) %>%
  group_by(`Pub ID`, response_first, response_second) %>%
  summarise(count = n(), .groups = "drop") %>%
  group_by(`Pub ID`) %>%
  filter(all(response_first == "No response")) %>%
  pivot_wider(
    values_from = count,
    names_from = response_second,
    id_cols = `Pub ID`,
    values_fill = 0
  ) %>%
  mutate(
    response_per_study = case_when(
      Response >= 1 ~ "Response",
      Failed >= 1 & Response == 0 & `No response` == 0 ~ "Failed",
      TRUE ~ "No response"
    )
  )

# Summarize response counts
failed_count <- sum(second_inquiry_responses$Failed)
response_count <- sum(second_inquiry_responses$Response)
no_response_count <- sum(second_inquiry_responses$`No response`)

paste("Failed count:", failed_count)
paste("Response count:", response_count)
paste("No response count:", no_response_count)

# Response counts on unique authors level
unique_author_responses <- data %>%
  filter(`Pub ID` %in% as.vector(t(responses_first_inquiry %>% filter(response_per_study == "No response") %>% select(`Pub ID`)))) %>% 
  filter(`Date of 1st inquiry via mail` != "N/A") %>%
  distinct(`Name of author`, .keep_all = TRUE) %>%
  group_by(`Response to 2nd inquiry via mail`) %>%
  summarise(Count = n())

failed_count_unique <- unique_author_responses$Count[unique_author_responses$`Response to 2nd inquiry via mail` == "Mail address not available anymore"] 
response_count_unique <- unique_author_responses$Count[unique_author_responses$`Response to 2nd inquiry via mail` == "Pending response"] +
  unique_author_responses$Count[unique_author_responses$`Response to 2nd inquiry via mail` == "Redirect/Referral response"] + 
  unique_author_responses$Count[unique_author_responses$`Response to 2nd inquiry via mail` == "Data sharing declined"] 
no_response_count_unique <- unique_author_responses$Count[unique_author_responses$`Response to 2nd inquiry via mail` == "No response"]  

paste("Failed count (unique authors):", failed_count_unique)
paste("Response count (unique authors):", response_count_unique)
paste("No response count (unique authors):", no_response_count_unique)

# Study-level response counts
study_response_counts <- second_inquiry_responses %>%
  group_by(response_per_study) %>%
  summarise(count = n())

paste("Failed count (study-level):", study_response_counts$count[study_response_counts$response_per_study == "Failed"])
paste("Response count (study-level):", study_response_counts$count[study_response_counts$response_per_study == "Response"])
paste("No response count (study-level):", study_response_counts$count[study_response_counts$response_per_study == "No response"])

### ResearchGate inquiry
# -------------------------------------

# number of studies eligible for researchgate contact
studies_eligible_ResearchGate <- data %>%
  filter(`Pub ID` %in% c(
    as.vector(t(second_inquiry_responses %>% filter(response_per_study == "No response" | response_per_study == "Failed") %>% select(`Pub ID`))), 
    as.vector(t(responses_first_inquiry %>% filter(response_per_study == "Failed") %>% select(`Pub ID`))))) %>%
  distinct(`Pub ID`, .keep_all = TRUE) %>%
  nrow()

paste("Number of studies eligible for contact via Researchgate:", studies_eligible_ResearchGate)

# number of authors eligible for ResearchGate contact
# including first authors (not previously contacted)
authors_eligible_ResearchGate <- data %>%
  filter(`Pub ID` %in% c(
    as.vector(t(second_inquiry_responses %>% filter(response_per_study == "No response" | response_per_study == "Failed") %>% select(`Pub ID`))), 
    as.vector(t(responses_first_inquiry %>% filter(response_per_study == "Failed") %>% select(`Pub ID`))))) %>%
  nrow()

paste("Number of authors eligible for contact via Researchgate:", authors_eligible_ResearchGate)

# Authors eligible for ResearchGate contact distinguished between
# previously contacted and not contacted
researchgate_contacts_author <- data %>%
  filter(`Pub ID` %in% c(
    as.vector(t(second_inquiry_responses %>% filter(response_per_study == "No response" | response_per_study == "Failed") %>% select(`Pub ID`))), 
    as.vector(t(responses_first_inquiry %>% filter(response_per_study == "Failed") %>% select(`Pub ID`))))) %>%
  mutate(
    prev_email_contact = ifelse(`Date of 1st inquiry via mail` != "N/A", TRUE, FALSE)
  ) %>%
  group_by(prev_email_contact, `Researchgate account available`) %>%
  summarise(count = n())

paste("Number of authors previously contacted without success:", sum(researchgate_contacts_author$count[researchgate_contacts_author$prev_email_contact == "TRUE"]))
paste("Number of authors previously contacted without success with account:", sum(researchgate_contacts_author$count[researchgate_contacts_author$prev_email_contact == "TRUE" & researchgate_contacts_author$`Researchgate account available` == "yes"]))

paste("Number of authors noot previously contacted eligible for ResearchGate contact:", sum(researchgate_contacts_author$count[researchgate_contacts_author$prev_email_contact == "FALSE"]))
paste("Number of authors previously contacted without success with account:", sum(researchgate_contacts_author$count[researchgate_contacts_author$prev_email_contact == "FALSE" & researchgate_contacts_author$`Researchgate account available` == "yes"]))

paste("Total number of authors eligible for ResearchGate contact with account:", sum(researchgate_contacts_author$count[researchgate_contacts_author$`Researchgate account available` == "yes"]))

# Researchgate contact per study
researchgate_per_study <- data %>%
  filter(`Pub ID` %in% c(
    as.vector(t(second_inquiry_responses %>% filter(response_per_study == "No response" | response_per_study == "Failed") %>% select(`Pub ID`))), 
    as.vector(t(responses_first_inquiry %>% filter(response_per_study == "Failed") %>% select(`Pub ID`))))) %>%
  mutate(
    prev_email_contact = ifelse(`Date of 1st inquiry via mail` != "N/A", TRUE, FALSE)
  ) %>%
  mutate(researchgate_contacted = ifelse(`Date of inquiry via Researchgate` != "N/A", TRUE, FALSE)) %>%
  group_by(`Pub ID`, researchgate_contacted) %>%
  summarise(
    contacted_via_researchgate = n()
  )

# Summarize contact counts
contacted_count <- sum(researchgate_per_study$researchgate_contacted)
not_contacted_count <- 34- sum(researchgate_per_study$researchgate_contacted)

print(paste("Studies contacted count:", contacted_count))
print(paste("Studies not contacted count:", not_contacted_count))

# Responses to Researchgate inquiry on author-level
responses_researchgate <- data %>%
  filter(`Pub ID` != 45) %>%
  group_by(`Pub ID`) %>%
  # Filter out publications with at least one response
  filter(!(any(`Response to 1st inquiry via mail` == "Data sharing declined") | 
             any(`Response to 1st inquiry via mail` == "Redirect/Referral response") | 
             any(`Response to 1st inquiry via mail` == "Pending response") | 
             any(`Response to 2nd inquiry via mail` == "Data sharing declined") |
             any(`Response to 2nd inquiry via mail` == "Redirect/Referral response") | 
             any(`Response to 2nd inquiry via mail` == "Pending response")),
         `Date of inquiry via Researchgate` != "N/A") %>%
  group_by(`Response to Researchgate inquiry`) %>%
  summarise(count = n())

paste("Number of contact attempts with response:", responses_researchgate$count[responses_researchgate$`Response to Researchgate inquiry` == "Data sharing declined"])
paste("Number of contact attempts without response:", responses_researchgate$count[responses_researchgate$`Response to Researchgate inquiry` == "No response"])

# Responses on unique authors level
responses_researchgate_unique_author <- data %>%
  filter(`Pub ID` != 45) %>%
  group_by(`Pub ID`) %>%
  # Filter out publications with at least one response
  filter(!(any(`Response to 1st inquiry via mail` == "Data sharing declined") | 
             any(`Response to 1st inquiry via mail` == "Redirect/Referral response") | 
             any(`Response to 1st inquiry via mail` == "Pending response") | 
             any(`Response to 2nd inquiry via mail` == "Data sharing declined") |
             any(`Response to 2nd inquiry via mail` == "Redirect/Referral response") | 
             any(`Response to 2nd inquiry via mail` == "Pending response")),
         `Date of inquiry via Researchgate` != "N/A") %>%
  group_by(`Response to Researchgate inquiry`) %>%
  distinct(`Name of author`, .keep_all = TRUE) %>%
  summarise(count = n())
paste("Number of unique authors with response:", responses_researchgate_unique_author$count[responses_researchgate_unique_author$`Response to Researchgate inquiry` == "Data sharing declined"])
paste("Number of unique authors without response:", responses_researchgate_unique_author$count[responses_researchgate_unique_author$`Response to Researchgate inquiry` == "No response"])

# Responses per study
responses_researchgate_per_study <- data %>%
  filter(`Pub ID` != 45) %>%
  group_by(`Pub ID`) %>%
  # Filter out publications with at least one response
  filter(!(any(`Response to 1st inquiry via mail` == "Data sharing declined") | 
             any(`Response to 1st inquiry via mail` == "Redirect/Referral response") | 
             any(`Response to 1st inquiry via mail` == "Pending response") | 
             any(`Response to 2nd inquiry via mail` == "Data sharing declined") |
             any(`Response to 2nd inquiry via mail` == "Redirect/Referral response") | 
             any(`Response to 2nd inquiry via mail` == "Pending response")),
         `Date of inquiry via Researchgate` != "N/A") %>%
  group_by(`Pub ID`, `Response to Researchgate inquiry`) %>%
  summarise(count = n()) %>%
  pivot_wider(
    values_from = count,
    names_from = `Response to Researchgate inquiry`,
    id_cols = `Pub ID`,
    values_fill = 0
  )

# Summarize response counts
no_response_count <- sum(responses_researchgate_per_study$`No response`[responses_researchgate_per_study$`Data sharing declined` == 0] >= 1)
print(paste("No response count (study-level):", no_response_count))

### Contacts and responses in total
# -------------------------------------

# total number of contacts
total_contact_count <- data %>%
  filter(`Pub ID` != 45) %>%
  pull(`Name of author`)
print(paste("Total number of contacted authors:", length(total_contact_count)))
print(paste("Total number of (unique) contacted authors:", length(unique(total_contact_count))))

# Responses per study on author-level
response_total_per_study <- data %>%
  filter(`Pub ID` != 45) %>%
  mutate(
    response_first = case_when(
      `Response to 1st inquiry via mail` == "Mail address not available anymore" ~ "Failed",
      `Response to 1st inquiry via mail` == "No response" ~ "No response",
      `Response to 1st inquiry via mail` == "N/A" ~ "N/A",
      TRUE ~ "Response"
    ),
    response_second = case_when(
      `Response to 2nd inquiry via mail` == "Mail address not available anymore" ~ "Failed",
      `Response to 2nd inquiry via mail` == "No response" ~ "No response",
      `Response to 2nd inquiry via mail` == "N/A" ~ "N/A",
      TRUE ~ "Response"
    ),
    response_researchgate = case_when(
      `Response to Researchgate inquiry` == "No response" ~ "No response",
      `Response to Researchgate inquiry` == "N/A" ~ "N/A",
      TRUE ~ "Response"
    )
  ) %>%
  mutate(
    response_per_author = case_when(
      response_first == "Response" | response_second == "Response" | response_researchgate == "Response" ~ TRUE,
      TRUE ~ FALSE
    )
  ) %>%
  group_by(`Pub ID`, response_per_author) %>%
  summarise(count = n()) %>%
  pivot_wider(
    values_from = count,
    names_from = response_per_author,
    id_cols = `Pub ID`,
    values_fill = 0
  )

# Summarize count of responsive authors
response_count_authors <- sum(response_total_per_study$`TRUE`)
no_response_count_authors <- sum(response_total_per_study$`FALSE`)

print(paste("Count of responsive authors:", response_count_authors))
print(paste("Count of unresponsive authors:", no_response_count_authors))

# Summarize count of responsive unique authors
response_total_per_study_unique_authors <- data %>%
  filter(`Pub ID` != 45) %>%
  mutate(
    response_first = case_when(
      `Response to 1st inquiry via mail` == "Mail address not available anymore" ~ "Failed",
      `Response to 1st inquiry via mail` == "No response" ~ "No response",
      `Response to 1st inquiry via mail` == "N/A" ~ "N/A",
      TRUE ~ "Response"
    ),
    response_second = case_when(
      `Response to 2nd inquiry via mail` == "Mail address not available anymore" ~ "Failed",
      `Response to 2nd inquiry via mail` == "No response" ~ "No response",
      `Response to 2nd inquiry via mail` == "N/A" ~ "N/A",
      TRUE ~ "Response"
    ),
    response_researchgate = case_when(
      `Response to Researchgate inquiry` == "No response" ~ "No response",
      `Response to Researchgate inquiry` == "N/A" ~ "N/A",
      TRUE ~ "Response"
    )
  ) %>%
  mutate(
    response_per_author = case_when(
      response_first == "Response" | response_second == "Response" | response_researchgate == "Response" ~ TRUE,
      TRUE ~ FALSE
    )
  ) %>%
  group_by(response_per_author) %>%
  distinct(`Name of author`, .keep_all = T) %>%
  summarise(count = n()) 

paste("Number of unique authors without response:", response_total_per_study_unique_authors$count[response_total_per_study_unique_authors$response_per_author == "FALSE"] )
paste("Number of unique authors with response:", response_total_per_study_unique_authors$count[response_total_per_study_unique_authors$response_per_author == "TRUE"] )


# Summarize count of responsive studies
response_count <- sum(response_total_per_study$`TRUE` >= 1)
no_response_count <- sum(response_total_per_study$`TRUE` == 0)

print(paste("Count of responsive studies:", response_count))
print(paste("Count of unresponsive studies:", no_response_count))


### Content of responses
# ------------------------------------

# Analyze responses
responses <- data %>%
  filter(`Pub ID` != 45) %>%
  mutate(
    response_first = case_when(
      `Response to 1st inquiry via mail` == "Mail address not available anymore" ~ "Failed",
      `Response to 1st inquiry via mail` == "No response" ~ "No response",
      `Response to 1st inquiry via mail` == "N/A" ~ "N/A",
      TRUE ~ "Response"
    ),
    response_second = case_when(
      `Response to 2nd inquiry via mail` == "Mail address not available anymore" ~ "Failed",
      `Response to 2nd inquiry via mail` == "No response" ~ "No response",
      `Response to 2nd inquiry via mail` == "N/A" ~ "N/A",
      TRUE ~ "Response"
    ),
    response_researchgate = case_when(
      `Response to Researchgate inquiry` == "No response" ~ "No response",
      `Response to Researchgate inquiry` == "N/A" ~ "N/A",
      TRUE ~ "Response"
    )
  ) %>%
  mutate(
    response_per_author = case_when(
      response_first == "Response" | response_second == "Response" | response_researchgate == "Response" ~ TRUE,
      TRUE ~ FALSE
    )
  ) %>%
  filter(response_per_author == TRUE) %>%
  mutate(
    response = case_when(
      `Response to 1st inquiry via mail` == "Data sharing declined" | 
        `Response to 1st inquiry via mail` == "Pending response" | 
        `Response to 1st inquiry via mail` == "Redirect/Referral response" ~ `Response to 1st inquiry via mail`,
      `Response to 2nd inquiry via mail` == "Data sharing declined" | 
        `Response to 2nd inquiry via mail` == "Redirect/Referral response" | 
        `Response to 2nd inquiry via mail` == "Pending response" ~ `Response to 2nd inquiry via mail`,
      `Response to Researchgate inquiry` == "Data sharing declined" | 
        `Response to Researchgate inquiry` == "Pending response" | 
        `Response to Researchgate inquiry` == "Redirect/Referral response" ~ `Response to Researchgate inquiry`
    )
  ) %>%
  group_by(`Pub ID`, `Name of author`, response) %>%
  summarise(count = n())

# Analyze response counts
response_counts <- responses %>%
  group_by(response) %>%
  summarise(count = n())

paste("Number of contacted authors with pending response:", response_counts$count[response_counts$response == "Pending response"])
paste("Number of contacted authors with redirect response:", response_counts$count[response_counts$response == "Redirect/Referral response"])
paste("Number of contacted authors response declining to share data:", response_counts$count[response_counts$response == "Data sharing declined"])

# Reasoning and action upon responses 
responses_reasoning_and_action <- data %>%
  filter(`Pub ID` != 45) %>%
  mutate(
    response_first = case_when(
      `Response to 1st inquiry via mail` == "Mail address not available anymore" ~ "Failed",
      `Response to 1st inquiry via mail` == "No response" ~ "No response",
      `Response to 1st inquiry via mail` == "N/A" ~ "N/A",
      TRUE ~ "Response"
    ),
    response_second = case_when(
      `Response to 2nd inquiry via mail` == "Mail address not available anymore" ~ "Failed",
      `Response to 2nd inquiry via mail` == "No response" ~ "No response",
      `Response to 2nd inquiry via mail` == "N/A" ~ "N/A",
      TRUE ~ "Response"
    ),
    response_researchgate = case_when(
      `Response to Researchgate inquiry` == "No response" ~ "No response",
      `Response to Researchgate inquiry` == "N/A" ~ "N/A",
      TRUE ~ "Response"
    )
  ) %>%
  mutate(
    response_per_author = case_when(
      response_first == "Response" | response_second == "Response" | response_researchgate == "Response" ~ TRUE,
      TRUE ~ FALSE
    )
  ) %>%
  filter(response_per_author == TRUE) %>%
  mutate(
    response = case_when(
      `Response to 1st inquiry via mail` == "Data sharing declined" | 
        `Response to 1st inquiry via mail` == "Pending response" | 
        `Response to 1st inquiry via mail` == "Redirect/Referral response" ~ `Response to 1st inquiry via mail`,
      `Response to 2nd inquiry via mail` == "Data sharing declined" | 
        `Response to 2nd inquiry via mail` == "Redirect/Referral response" | 
        `Response to 2nd inquiry via mail` == "Pending response" ~ `Response to 2nd inquiry via mail`,
      `Response to Researchgate inquiry` == "Data sharing declined" | 
        `Response to Researchgate inquiry` == "Pending response" | 
        `Response to Researchgate inquiry` == "Redirect/Referral response" ~ `Response to Researchgate inquiry`
    )
  ) %>%
  select(`Pub ID`, response, `Reasoning of author/Content of response`, `Action upon response`)
print(responses_reasoning_and_action)
```
