## Supplement 4: R Source code for analysis and creation of graphics for "A Case Study on Colposcopy-Based Cervical Cancer Staging Reveals an Alarming Lack of Data Sharing Hindering the Adoption of Machine Learning in Clinical Practice": 3.4_Factors influencing data sharing and response behavior.R

```
# setwd() # set appropriate working directory

library(readxl)
library(dplyr)
library(tidyverse)

data <- read_xlsx("Supplement3_Detailed_results.xlsx", sheet = "Data inquiry details")
DAS_info <- read_xlsx("Supplement3_Detailed_results.xlsx", sheet = "analysis_table")

### data preparation
# -------------------------------------

responses_with_das <- data %>%
  filter(`Pub ID` != 45) %>%
  mutate(
    response_first = case_when(
      `Response to 1st inquiry via mail` == "Mail address not available anymore" ~ "Failed",
      `Response to 1st inquiry via mail` == "No response" ~ "No response",
      `Response to 1st inquiry via mail` == "N/A" ~ "N/A",
      TRUE ~ "Response"
    ),
    response_second = case_when(
      `Response to 2nd inquiry via mail` == "Mail address not available anymore" ~ "Failed",
      `Response to 2nd inquiry via mail` == "No response" ~ "No response",
      `Response to 2nd inquiry via mail` == "N/A" ~ "N/A",
      TRUE ~ "Response"
    ),
    response_researchgate = case_when(
      `Response to Researchgate inquiry` == "No response" ~ "No response",
      `Response to Researchgate inquiry` == "N/A" ~ "N/A",
      TRUE ~ "Response"
    )
  ) %>%
  mutate(
    response_per_author = case_when(
      response_first == "Response" | response_second == "Response" | response_researchgate == "Response" ~ "Response",
      TRUE ~ "No Response"
    )
  ) %>%
  group_by(`Pub ID`,response_per_author) %>%
  summarise(count = n()) %>%
  pivot_wider(
    values_from = count,
    names_from = response_per_author,
    id_cols = `Pub ID`,
    values_fill = 0
  ) %>%
  mutate(
    response_per_study = case_when(
      Response >= 1 ~ "Response",
      TRUE ~ "No response"
    )
  ) %>%
  merge(DAS_info, by = "Pub ID") %>%
  mutate(DAS = case_when(`DAS available (Y/N)` == "N"  ~ "DAS unavailable",
                         `DAS available (Y/N)` == "Y" & `Data declared available (Y/N)` == "Y" ~ "Positive DAS",
                         `DAS available (Y/N)` == "Y" & `Data declared available (Y/N)` == "N" ~ "Negative DAS"
  )) 

# numbers Figure 3, data sharing outcome by status of DAS
dataavailab_das <- data %>%
  filter(`Pub ID` != 45) %>%
  group_by(`Pub ID`, Outcome) %>%
  summarise(count = n()) %>%
  pivot_wider(
    values_from = count,
    names_from = Outcome,
    id_cols = `Pub ID`,
    values_fill = 0
  ) %>%
  mutate(
    dataavailab_per_study = ifelse(`Data not shared` >= 1, "Data not shared", "Data shared")
  ) %>%
  merge(DAS_info, by = "Pub ID") %>%
  mutate(DAS = case_when(`DAS available (Y/N)` == "N"  ~ "DAS unavailable",
                         `DAS available (Y/N)` == "Y" & `Data declared available (Y/N)` == "Y" ~ "Positive DAS",
                         `DAS available (Y/N)` == "Y" & `Data declared available (Y/N)` == "N" ~ "Negative DAS"
  )) %>%
  group_by(dataavailab_per_study, DAS) %>%
  summarise(count = n()) %>%
  rbind(data.frame(dataavailab_per_study = "Data shared", DAS = "DAS unavailable", count = 0))
dataavailab_das

# numbers Figure 3, responses received by status of DAS
responses_das_analysis <- responses_with_das %>%
  group_by(response_per_study, DAS) %>%
  summarise(count = n()) 
responses_das_analysis

### Analysis I: Status DAS
# -------------------------------------

responses_das_status <- responses_with_das %>%
  group_by(response_per_study, `DAS available (Y/N)`) %>%
  summarise(count = n()) 

# frequency table
table <- xtabs(count ~ response_per_study + `DAS available (Y/N)`, data = responses_das_status)
# results of chi2 test of independence
DAS_avail_df <- data.frame(variable = "DAS availability", chisq.test(table, simulate.p.value = T)[c("statistic", "p.value")], row.names = NULL)
print(DAS_avail_df)

### Analysis II: Content DAS
# -------------------------------------

# frequency table
table <- xtabs(count ~ response_per_study + DAS, data = responses_das_analysis[responses_das_analysis$DAS != "DAS unavailable",])
# results of chi2 test of independence
DAS_df <- data.frame(variable = "DAS", chisq.test(table, simulate.p.value = T)[c("statistic", "p.value")], row.names = NULL)
print(DAS_df)

### Analysis III: Region
# -------------------------------------

continent_mapping <- c("USA" = "North America",
                       "China" = "Asia",
                       "Korea" = "Asia",
                       "Japan" = "Asia",
                       "India" = "Asia",
                       "Netherlands" = "Europe",
                       "Costa Rica" = "North America",
                       "France" = "Europe")
responses_with_das$continent_publication <- continent_mapping[responses_with_das$`Primary country of publication origin`]

responses_region_analysis <- responses_with_das %>%
  group_by(response_per_study, continent_publication) %>%
  summarise(count = n()) 

table <- xtabs(count ~ response_per_study +  continent_publication, data = responses_region_analysis)
region_df <- data.frame(variable = "Region", chisq.test(table, simulate.p.value = T)[c("statistic",  "p.value")], row.names = NULL)

### Analysis IV: Year (Difference)
# -------------------------------------

responses_with_das$year_diff <- 2024-responses_with_das$Year
responses_with_das$response_per_study <- factor(responses_with_das$response_per_study)
levels(responses_with_das$response_per_study) <- c(0,1)

# mean and standard deviation by response:
responses_with_das %>% group_by(response_per_study) %>% summarise(mean = mean(year_diff), sd = sd(year_diff))

log_year <- glm(response_per_study ~ year_diff, data = responses_with_das, family = binomial(link = "logit"))
summary(log_year)

### Analysis V: Number of image data
# -------------------------------------

# mean and standard deviation by response:
responses_with_das %>% group_by(response_per_study) %>% summarise(mean = mean(`Total number of image data`), sd = sd(`Total number of image data`))

log_numberofimagedata <- glm(response_per_study ~ `Total number of image data`, data = responses_with_das, family = binomial(link = "logit"))
summary(log_numberofimagedata)

### Analysis VI: AUC
# -------------------------------------

responses_with_das$`AUC: Binary classification_1` <- as.numeric(as.character(responses_with_das$`AUC: Binary classification_1`))
responses_with_das$`AUC: Binary classification_2` <- as.numeric(as.character(responses_with_das$`AUC: Binary classification_2`))
responses_with_das$`AUC: Multiclass classification` <- as.numeric(as.character(responses_with_das$`AUC: Multiclass classification`))

# Create a new variable with the best performance metrics for each model
responses_with_das$best_auc <- apply(responses_with_das[, c("AUC: Binary classification_1", "AUC: Binary classification_2", "AUC: Multiclass classification")], 1, function(x) {
  if (all(is.na(x))) {
    return(NA)
  } else {
    return(max(x, na.rm = TRUE))
  }
})

sum(!is.na(responses_with_das$best_auc))

# mean and standard deviation by response
responses_with_das %>% group_by(response_per_study) %>% summarise(mean = mean(best_auc, na.rm = T), sd = sd(best_auc, na.rm = T))

log_performance <- glm(response_per_study ~ best_auc, data = responses_with_das, family = binomial(link = "logit"))
summary(log_performance)

### Analysis VII: Accuracy
# -------------------------------------

responses_with_das$`Accuracy: Binary classification_1` <- as.numeric(as.character(responses_with_das$`Accuracy: Binary classification_1`))
responses_with_das$`Accuracy: Binary classification_2` <- as.numeric(as.character(responses_with_das$`Accuracy: Binary classification_2`))
responses_with_das$`Accuracy: Multiclass classification` <- as.numeric(as.character(responses_with_das$`Accuracy: Multiclass classification`))

# Create a new variable with the best performance metrics for each model
responses_with_das$best_accuracy <- apply(responses_with_das[, c("Accuracy: Binary classification_1", "Accuracy: Binary classification_2", "Accuracy: Multiclass classification")], 1, function(x) {
  if (all(is.na(x))) {
    return(NA)
  } else {
    return(max(x, na.rm = TRUE))
  }
})

sum(!is.na(responses_with_das$best_accuracy))

# mean and standard deviation by response
responses_with_das %>% group_by(response_per_study) %>% summarise(mean = mean(best_accuracy, na.rm = T), sd = sd(best_accuracy, na.rm = T))

log_performance2 <- glm(response_per_study ~ best_accuracy, data = responses_with_das, family = binomial(link = "logit"))
summary(log_performance2)
```
