## Supplement 4: R Source code for analysis and creation of graphics for "A Case Study on Colposcopy-Based Cervical Cancer Staging Reveals an Alarming Lack of Data Sharing Hindering the Adoption of Machine Learning in Clinical Practice": Appendix_B.R

```
# setwd() # set appropriate working directory

library(readxl)
library(dplyr)
library(tidyverse)

data <- read_xlsx("Supplement3_Detailed_results.xlsx", sheet = "analysis_table" )

### B.1 Performed Tasks
# -----------------------------------

task_type <- data %>%
  filter(`Type of publication`== "Analysis publication") %>%
  group_by(`Task type`) %>%
  summarise(Count = n()) 

paste("Number of studies performing classification tasks:", task_type$Count[task_type$`Task type` == "Classification"])
paste("Number of studies performing segmentation tasks:", task_type$Count[task_type$`Task type` == "Segmentation"])
paste("Number of studies performing both segmentation and classification tasks:", task_type$Count[task_type$`Task type` == "Segmentation/Classification"])

classification_task <- data %>% 
  filter(`Type of publication`== "Analysis publication",
         `Task type` != "Segmentation") %>%
  group_by(`Classification task type`) %>%
  summarise(Count = n()) 

paste("Number of studies performing binary classification:", classification_task$Count[classification_task$`Classification task type` == "Binary"])
paste("Number of studies performing multiclass classification:", classification_task$Count[classification_task$`Classification task type` == "Multiclass"])
paste("Number of studies performing binary and multiclass classification:", classification_task$Count[classification_task$`Classification task type` == "Multiclass + Binary"])

classification_grading <- data %>% 
  filter(`Type of publication`== "Analysis publication",
         `Task type` != "Segmentation") %>%
  group_by(`Grading system`) %>%
  summarise(Count = n()) 

paste("Number of studies usign WHO system:", classification_grading$Count[classification_grading$`Grading system` == "WHO"])
paste("Number of studies usign CIN system:", classification_grading$Count[classification_grading$`Grading system` == "CIN"])
paste("Number of studies unclear grading system:", classification_grading$Count[classification_grading$`Grading system` == "not given"])

binary_classification_details <- data %>% 
  filter(`Type of publication` == "Analysis publication",
         `Task type` != "Segmentation",
         `Classification task type` != "Multiclass") %>%
  group_by(`Binary classification details`) %>%
  summarise(Count =n())
print(binary_classification_details)

multiclass_classification_details <- data %>% 
  filter(`Type of publication` == "Analysis publication", 
                `Task type` != "Segmentation", 
                `Classification task type` != "Binary") %>%
  group_by(`Multiclass classification details`) %>%
  summarise(Count =n())
print(multiclass_classification_details)

### B.3 Details on the data used
# -------------------------------------

data$`Country of data collection`[data$`Country of data collection`== "North America, Central America, South America, Africa, and Asia"] <- "Various"
data$`Country of data collection`[data$`Country of data collection`== "USA, Peru, Tanzania, Zambia"] <- "Various"

ordered_countries <- c("China", "Korea", "Japan", "India", "Nigeria", "Zambia", "Netherlands", "Spain, Netherlands", "Costa Rica", "USA", "Various")

data_region <- data %>% 
  filter(`Use of external data sources (Y/N)` != "Y",
                `Type of publication` != "Publicly available dataset") %>%
  group_by(`Country of data collection`) %>%
  summarise(Count = n())
print(data_region)

# number of image data
summary(data %>% 
          filter(`Type of publication` == "Analysis publication") %>%
          select(`Total number of image data`))

# studies validating pre-trained models: Pub ID 6, 18, 19, 34, 35
summary(data %>% 
          filter(`Type of publication` == "Analysis publication",
                 `Pub ID`== 6 | `Pub ID`== 18 | `Pub ID`== 19 |`Pub ID`== 34 |`Pub ID`== 35) %>%
          select(`Total number of image data`))

# without validation studies
summary(data %>% 
          filter(`Type of publication` == "Analysis publication",
                 `Pub ID`!= 6 , `Pub ID`!= 18 , `Pub ID`!= 19 ,`Pub ID`!= 34 ,`Pub ID`!= 35) %>%
          select(`Total number of image data`))

# Class distribution (binary tasks)
class_distr <- data$`Class distribution of image data for binary classification`
class_distr <- subset(class_distr, !(class_distr %in% c("N/A", "not reported")))
class_distr <- as.numeric(class_distr)

summary(class_distr)*100
boxplot(class_distr)

### B.4 Reported performance measures
# -------------------------------------

# AUC Binary
# Number of studies AUC not reported:
data %>% filter(`Task type` != "Segmentation", `Type of publication` == "Analysis publication", `Classification task type` != "Multiclass") %>% group_by(`AUC: Binary classification_1`== "not reported") %>% summarise(Count = n())

auc_binary <- c(data$`AUC: Binary classification_1`, data$`AUC: Binary classification_2`)
auc_binary <- subset(auc_binary, !(auc_binary %in% c("N/A", "not reported")))
auc_binary <- as.numeric(auc_binary)

summary(auc_binary)

# Accuracy Binary
# accuracy not reported:
data %>% filter(`Task type` != "Segmentation", `Type of publication` == "Analysis publication", `Classification task type` != "Multiclass") %>% group_by(`Accuracy: Binary classification_1`== "not reported") %>% summarise(Count = n())

acc_binary <- c(data$`Accuracy: Binary classification_1`, data$`Accuracy: Binary classification_2`)
acc_binary <- subset(acc_binary, !(acc_binary %in% c("N/A", "not reported")))
acc_binary <- as.numeric(acc_binary)

summary(acc_binary)

# AUC Multiclass
# not reported:
data %>% filter(`Task type` != "Segmentation", `Type of publication` == "Analysis publication", `Classification task type` != "Binary") %>% group_by(`AUC: Multiclass classification`== "not reported") %>% summarise(Count = n())

auc_multi <- c(data$`AUC: Multiclass classification`)
auc_multi <- subset(auc_multi, !(auc_multi %in% c("N/A", "not reported")))
auc_multi <- as.numeric(auc_multi)

summary(auc_multi)

# Accuracy Multiclass
# not reported: 
data %>% filter(`Task type` != "Segmentation", `Type of publication` == "Analysis publication", `Classification task type` != "Binary") %>% group_by(`Accuracy: Multiclass classification`== "not reported") %>% summarise(Count = n())

acc_multiclass <- c(data$`Accuracy: Multiclass classification`)
acc_multiclass <- subset(acc_multiclass, !(acc_multiclass %in% c("N/A", "not reported")))
acc_multiclass <- as.numeric(acc_multiclass)

summary(acc_multiclass)
```
