## Supplement 4: R Source code for analysis and creation of graphics for "A Case Study on Colposcopy-Based Cervical Cancer Staging Reveals an Alarming Lack of Data Sharing Hindering the Adoption of Machine Learning in Clinical Practice": graphics.R

```
# setwd() # set appropriate working directory

library(readxl)
library(dplyr)
library(tidyverse)
library(ggplot2)
library(ggpubr)
library(gridExtra)

data <- read_xlsx("Supplement3_Detailed_results.xlsx", sheet = "analysis_table" )

###########################
###### FIGURE 1 ###########
###########################


### DAS available
# --------------------------------------

das_available <- data %>% filter(`Type of publication` != "Publicly available dataset") %>%
  mutate(type = case_when(`Use of external data sources (Y/N)`== "Y" ~ "Analysis publication using external data source",
                          `Use of external data sources (Y/N)`== "N" ~ "Analysis publication using own dataset",
                          `Type of publication` == "Publication providing data" ~ "Publication providing data")) %>%
  group_by(type, `DAS available (Y/N)`) %>%
  summarise(Count = n()) %>%
  ggplot(aes(fill=type, y=Count, x=`DAS available (Y/N)`)) + 
  geom_bar(position="stack", stat="identity") +
  geom_text(aes(label=Count), position=position_stack(vjust=0.5), size = 6) +
  labs(subtitle = "DAS Availability", y="Number of studies", fill ="", x = "") +
  guides(fill=guide_legend(nrow=3, byrow=TRUE))+
  scale_x_discrete(labels=c("N" = "No DAS included", "Y" = "DAS included")) + 
  # scale_fill_brewer(palette = "YlGnBu")+
  scale_fill_manual(values = c("#00AFBB", "#EEA236", "#8F3931"))+
  theme_minimal() +
  theme(legend.position = "top",
        plot.title = element_text(hjust = 0.5, size = 15, margin = margin(b = 5), face="bold"),
        plot.subtitle = element_text(hjust = 0, size = 20, margin = margin(b = 10)),
        panel.border = element_rect(color = "grey90", fill = NA, linewidth = 1),
        axis.title.x = element_text(size = 15, margin = margin(t = 10)),
        axis.text=element_text(size=15),
        legend.text=element_text(size=15),
        axis.title.y = element_text(size = 15, margin = margin(r = 10), vjust = 0.5),
        plot.caption = element_text(size = 5, margin = margin(t = 10), hjust = 0),
        legend.text.align = 0
  )

### DAS over time
#---------------------------------------

data_sum <- data %>%
  filter(`Type of publication` != "Publicly available dataset") %>%
  mutate(Year2 = ifelse(Year < 2018, "<2018", as.character(Year))) %>%
  group_by(Year2) %>%
  summarise(
    Count_Y = sum(`DAS available (Y/N)` == "Y"),
    Total_Count = n()) %>%
  mutate(Proportion = (Count_Y / Total_Count)*100)

# Pivot data to longer format
data_long <- data_sum %>%
  pivot_longer(c(Count_Y, Total_Count), names_to = "variable", values_to = "value")


das_time <- ggplot(data_long, aes(x = Year2, y = value, fill = variable)) +
  geom_bar(stat = "identity", position = "dodge") +
  labs(subtitle = "DAS over time", x="Year of publication", y="Number of studies", fill ="") +
  geom_text(aes(x = Year2, y = value, label = ifelse(variable == "Count_Y", paste(round(Proportion), "%"), "")),
            position = position_dodge(width = .9), vjust = -1, size = 6) +
  # scale_fill_brewer(labels =c("Studies with DAS", "Included studies"), palette = "YlGnBu")+
  scale_fill_manual(values = c( "#EEA236", "#00AFBB","#8F3931"), labels =c("Studies with DAS", "Included studies"))+
  theme_minimal() +
  theme(legend.position = "top",
        plot.title = element_text(hjust = 0.5, size = 15, margin = margin(b = 5), face="bold"),
        plot.subtitle = element_text(hjust = 0, size = 20, margin = margin(b = 10)),
        panel.border = element_rect(color = "grey90", fill = NA, linewidth = 1),
        axis.title.x = element_text(size = 15, margin = margin(t = 10)),
        axis.text=element_text(size=15),
        legend.text=element_text(size=15),
        axis.title.y = element_text(size = 15, margin = margin(r = 10), vjust = 0.5),
        plot.caption = element_text(size = 5, margin = margin(t = 10), hjust = 0)
  )

### Generic DAS
# --------------------------------------

generic_das <- data %>% filter(`Type of publication` != "Publicly available dataset",
                `DAS available (Y/N)` == "Y", 
                # `Data declared available (Y/N)` == "Y"
                ) %>%
  group_by(`Generic DAS with data available on request`) %>%
  summarise(Count = n())

generic_das_plot <- ggplot(generic_das,
       aes(x = Count,
           xend = 0,
           y = `Generic DAS with data available on request`,
           yend = `Generic DAS with data available on request`,
           colour = `Generic DAS with data available on request`)) +
  geom_segment(linewidth = 5) +
  geom_point(size = 20)+ 
  scale_colour_manual(values = c("#00AFBB","#8F3931"), labels =c("Y" ="Generic DAS","N"= "Specific DAS"))+
  labs(subtitle = "Generic DAS", y = "", x = "Number of studies", col = "") +
  guides(col = "none") +
  scale_y_discrete(labels =c("Y" ="Generic DAS","N"= "Specific DAS"))+
  theme_void() +
  xlim(0,22)+
  geom_text(aes(label=Count), col = "black", position=position_stack(vjust=1), size = 6) +
  theme(legend.position = "top",
        plot.title = element_text(hjust = 0.5, size = 15, margin = margin(b = 5), face="bold"),
        plot.subtitle = element_text(hjust = 0, size = 20, margin = margin(b = 10, t = 5)),
        panel.border = element_rect(color = "grey90", fill = NA, linewidth = 1),
        axis.title.x = element_text(size = 15, margin = margin(t = 10)),
        axis.text=element_text(size=15),
        axis.text.y = element_text(angle = 80, hjust = 0.5),
        legend.text=element_text(size=15),
        axis.title.y = element_text(size = 15, margin = margin(r = 10), vjust = 0.5),
        plot.caption = element_text(size = 5, margin = margin(t = 10), hjust = 0)
  )

### Availability of what
# --------------------------------------

#raw data
DAS_rawdata <- data %>% filter(`Type of publication` != "Publicly available dataset",
                `DAS available (Y/N)` == "Y", 
                `Generic DAS with data available on request` == "N") %>%
  group_by(`Raw data available (Y/N)`) %>%
  summarise(Count = n()) 

new_row <- data.frame(`Raw data available (Y/N)` = "Not mentioned", Count = 0)
colnames(new_row) <- c("Raw data available (Y/N)", "Count")

DAS_rawdata <-  DAS_rawdata %>%
  rbind(new_row) %>%
  ggplot(aes(fill=`Raw data available (Y/N)`, y=Count, x=`Raw data available (Y/N)`)) + 
  geom_bar(position="stack", stat="identity") +
  geom_text(aes(label=Count), position=position_stack(vjust=0.5), size = 6) +
  labs(title = "Raw data", x="", y="", fill = "") +
  guides(fill = "none")+
  scale_x_discrete(label = c("N" = "Unavailable", "Y" = "Available", "Not mentioned" = "Not mentioned"),
                   limits = c("Not mentioned",  "N", "Y")) +
  ylim(0,22)+
  # scale_fill_brewer(palette = "YlGnBu")+
  scale_fill_manual(values = c( "#EEA236",  "#8F3931", "#00AFBB"))+
  theme_minimal() +
  theme(legend.position = "top",
        plot.subtitle = element_text(hjust = 0, size = 20, margin = margin(b = 10)),
        plot.title = element_text(hjust = 0, size = 15, margin = margin(b = 10)),
        panel.border = element_rect(color = "grey90", fill = NA, linewidth = 1),
        axis.title.x = element_text(size = 15, margin = margin(t = 10)),
        axis.text=element_text(size=15),
        legend.text=element_text(size=15),
        axis.title.y = element_text(size = 15, margin = margin(r = 10), vjust = 0.5),
        plot.caption = element_text(size = 12, margin = margin(t = 10), hjust = 0)
  ) +
  coord_flip()

data %>% filter(`Type of publication` != "Publicly available dataset",
                `DAS available (Y/N)` == "Y", `Data declared available (Y/N)` == "Y",
                `Generic DAS with data available on request` == "N", `Raw data available (Y/N)` == "Y") %>%
  group_by(`Accessibility of raw data`) %>%
  summarise(Count = n())

#code
DAS_code <- data %>% filter(`Type of publication` != "Publicly available dataset",
                `DAS available (Y/N)` == "Y", 
                # `Data declared available (Y/N)` == "Y",
                `Generic DAS with data available on request` == "N") %>%
  group_by(`Code available (Y/N)`) %>%
  summarise(Count = n()) 

new_row <- data.frame(`Code available (Y/N)` = "N", Count = 0)
colnames(new_row) <- c("Code available (Y/N)", "Count")

DAS_code <- DAS_code %>%
  rbind(new_row) %>%
  ggplot(aes(fill=`Code available (Y/N)`, y=Count, x=`Code available (Y/N)`)) + 
  geom_bar(position="stack", stat="identity") +
  geom_text(aes(label=Count), position=position_stack(vjust=0.5), size = 6) +
  labs(title = "Code", x="", y="", fill = "") +
  guides(fill = "none")+
  scale_x_discrete(label = c("N" = "Unavailable", "Y" = "Available", "Not mentioned" = "Not mentioned"),
                   limits = c("Not mentioned",  "N", "Y")) +
  ylim(0,22)+
  # scale_fill_brewer(palette = "YlGnBu")+
  scale_fill_manual(values = c( "#EEA236",  "#8F3931", "#00AFBB"))+
  theme_minimal() +
  theme(legend.position = "top",
        plot.subtitle = element_text(hjust = 0, size = 20, margin = margin(b = 10)),
        plot.title = element_text(hjust = 0, size = 15, margin = margin(b = 10)),
        panel.border = element_rect(color = "grey90", fill = NA, linewidth = 1),
        axis.title.x = element_text(size = 15, margin = margin(t = 10)),
        axis.text=element_text(size=15),
        legend.text=element_text(size=15),
        axis.title.y = element_text(size = 15, margin = margin(r = 10), vjust = 0.5),
        plot.caption = element_text(size = 12, margin = margin(t = 10), hjust = 0)
  ) + 
  coord_flip()

data %>% filter(`Type of publication` != "Publicly available dataset",
                `DAS available (Y/N)` == "Y", `Data declared available (Y/N)` == "Y",
                `Generic DAS with data available on request` == "N", `Code available (Y/N)` == "Y") %>%
  group_by(`Accessibility of code`) %>%
  summarise(Count = n())

#pre-trained weights
DAS_weights <- data %>% filter(`Type of publication` != "Publicly available dataset",
                `DAS available (Y/N)` == "Y", 
                # `Data declared available (Y/N)` == "Y",
                `Generic DAS with data available on request` == "N") %>%
  group_by(`Pre-trained weights available (Y/N)`) %>%
  summarise(Count = n()) 

new_row <- data.frame(`Pre-trained weights available (Y/N)` = c("Y", "N"), Count = c(0,0))
colnames(new_row) <- c("Pre-trained weights available (Y/N)", "Count")

DAS_weights <- DAS_weights %>%
  rbind(new_row) %>%
  ggplot(aes(fill=`Pre-trained weights available (Y/N)`, y=Count, x=`Pre-trained weights available (Y/N)`)) + 
  geom_bar(position="stack", stat="identity") +
  geom_text(aes(label=Count), position=position_stack(vjust=0.5), size = 6) +
  labs(x = "", title="Pre-trained weights", y="", fill = "") +
  guides(fill = "none")+
  scale_x_discrete(label = c("N" = "Unavailable", "Y" = "Available", "Not mentioned" = "Not mentioned"),
                   limits = c("Not mentioned",  "N", "Y")) +
  ylim(0,22)+
  # scale_fill_brewer(palette = "YlGnBu")+
  scale_fill_manual(values = c( "#EEA236",  "#8F3931", "#00AFBB"))+
  theme_minimal() +
  theme(legend.position = "top",
        plot.subtitle = element_text(hjust = 0, size = 20, margin = margin(b = 10)),
        plot.title = element_text(hjust = 0, size = 15, margin = margin(b = 10)),
        panel.border = element_rect(color = "grey90", fill = NA, linewidth = 1),
        axis.title.x = element_text(size = 15, margin = margin(t = 10)),
        axis.text=element_text(size=15),
        legend.text=element_text(size=15),
        axis.title.y = element_text(size = 15, margin = margin(r = 10), vjust = 0.5),
        plot.caption = element_text(size = 12, margin = margin(t = 10), hjust = 0)
  ) +
  coord_flip()


overall_combine <- ggarrange(ggarrange(das_available,ggplot() + theme_void(),das_time, nrow = 3, labels = c("A","", "B"), heights = c(1,0.05, 1)),
          ggplot() + theme_void(),
          ggarrange(
            generic_das_plot,
            ggplot() + theme_void(), 
            ggplot() + 
              labs(title = "Content of DAS") +
              theme(
                plot.title = element_text(hjust = 0, size = 20, margin = margin(b = 0)),
                plot.margin = margin(l = 40, t = 5),
                panel.background = element_blank()),
            ggarrange(DAS_rawdata, DAS_code, DAS_weights, nrow = 3),
            nrow = 4,
            heights = c(0.55,0.05,0.05, 1), labels = c("C", "", "D", "")),
          ncol = 3, widths = c(1,0.05,1))

ggsave(overall_combine,
    filename = "Figure1.pdf",
    width = 14,  # Adjust the width of the image
    height = 14,  # Adjust the height of the image
    dpi = 1000,  # Set the minimum dpi
    units = "in"
  )

###########################
###### FIGURE 3 ###########
###########################

# read in data -------------------------
data <- read_xlsx("Supplement3_Detailed_results.xlsx", sheet = "Data inquiry details")
DAS_info <- read_xlsx("Supplement3_Detailed_results.xlsx", sheet = "analysis_table" )

# prepare data -------------------------
responses_with_das <- data %>%
  filter(`Pub ID` != 45) %>%
  mutate(
    response_first = case_when(
      `Response to 1st inquiry via mail` == "Mail address not available anymore" ~ "Failed",
      `Response to 1st inquiry via mail` == "No response" ~ "No response",
      `Response to 1st inquiry via mail` == "N/A" ~ "N/A",
      TRUE ~ "Response"
    ),
    response_second = case_when(
      `Response to 2nd inquiry via mail` == "Mail address not available anymore" ~ "Failed",
      `Response to 2nd inquiry via mail` == "No response" ~ "No response",
      `Response to 2nd inquiry via mail` == "N/A" ~ "N/A",
      TRUE ~ "Response"
    ),
    response_researchgate = case_when(
      `Response to Researchgate inquiry` == "No response" ~ "No response",
      `Response to Researchgate inquiry` == "N/A" ~ "N/A",
      TRUE ~ "Response"
    )
  ) %>%
  mutate(
    response_per_author = case_when(
      response_first == "Response" | response_second == "Response" | response_researchgate == "Response" ~ "Response",
      TRUE ~ "No Response"
    )
  ) %>%
  group_by(`Pub ID`,response_per_author) %>%
  summarise(count = n()) %>%
  pivot_wider(
    values_from = count,
    names_from = response_per_author,
    id_cols = `Pub ID`,
    values_fill = 0
  ) %>%
  mutate(
    response_per_study = case_when(
      Response >= 1 ~ "Response",
      TRUE ~ "No response"
    )
  ) %>%
  merge(DAS_info, by = "Pub ID") %>%
  mutate(DAS = case_when(`DAS available (Y/N)` == "N"  ~ "DAS unavailable",
                         `DAS available (Y/N)` == "Y" & `Data declared available (Y/N)` == "Y" ~ "Positive DAS",
                         `DAS available (Y/N)` == "Y" & `Data declared available (Y/N)` == "N" ~ "Negative DAS"
  )) 

dataavailab_das <- data %>%
  filter(`Pub ID` != 45) %>%
  group_by(`Pub ID`, Outcome) %>%
  summarise(count = n()) %>%
  pivot_wider(
    values_from = count,
    names_from = Outcome,
    id_cols = `Pub ID`,
    values_fill = 0
  ) %>%
  mutate(
    dataavailab_per_study = ifelse(`Data not shared` >= 1, "Data not shared", "Data shared")
  ) %>%
  merge(DAS_info, by = "Pub ID") %>%
  mutate(DAS = case_when(`DAS available (Y/N)` == "N"  ~ "DAS unavailable",
                         `DAS available (Y/N)` == "Y" & `Data declared available (Y/N)` == "Y" ~ "Positive DAS",
                         `DAS available (Y/N)` == "Y" & `Data declared available (Y/N)` == "N" ~ "Negative DAS"
  )) %>%
  group_by(dataavailab_per_study, DAS) %>%
  summarise(count = n()) %>%
  rbind(data.frame(dataavailab_per_study = "Data shared", DAS = "DAS unavailable", count = 0))

responses_das_analysis <- responses_with_das %>%
  group_by(response_per_study, DAS) %>%
  summarise(count = n()) 

# plot-------------------------------
response_plot <- ggplot(responses_das_analysis, aes(x = response_per_study, y = count, fill = DAS)) +
  geom_bar(position="stack", stat="identity") +
  geom_text(aes(label=count), position=position_stack(vjust=0.5), size = 6) +
  labs(x="", y="Number of studies", fill ="") +
  # scale_fill_brewer(palette = "YlGnBu")+
  scale_fill_manual(values = c( "#EEA236", "#8F3931","#00AFBB"))+
  theme_minimal() +
  theme(
    legend.position = "top",
    plot.title = element_text(hjust = 0, size = 15, margin = margin(b = 5), face="bold"),
    plot.subtitle = element_text(hjust = 0, size = 20, margin = margin(b = 10)),
    panel.border = element_rect(color = "grey90", fill = NA, linewidth = 1),
    axis.title.x = element_text(size = 15, margin = margin(t = 10)),
    axis.text=element_text(size=15),
    legend.text=element_text(size=15),
    axis.title.y = element_text(size = 15, margin = margin(r = 10), vjust = 0.5),
    plot.caption = element_text(size = 5, margin = margin(t = 10), hjust = 0),
    legend.text.align = 0
  )

datasharing_plot <- ggplot(dataavailab_das, aes(x = dataavailab_per_study, y = count, fill = DAS)) +
  geom_bar(position = "stack", stat="identity") +
  geom_text(aes(label=count), position=position_stack(vjust=0.5), size = 6) +
  labs(x="", y="", fill ="") +
  # scale_fill_brewer(palette = "YlGnBu")+
  scale_fill_manual(values = c( "#EEA236", "#8F3931","#00AFBB"))+
  theme_minimal() +
  theme(
    legend.position = "top",
    plot.title = element_text(hjust = 0.5, size = 15, margin = margin(b = 5), face="bold"),
    plot.subtitle = element_text(hjust = 0, size = 20, margin = margin(b = 10)),
    panel.border = element_rect(color = "grey90", fill = NA, linewidth = 1),
    axis.title.x = element_text(size = 15, margin = margin(t = 10)),
    axis.text=element_text(size=15),
    legend.text=element_text(size=15),
    axis.title.y = element_text(size = 15, margin = margin(r = 10), vjust = 0.5),
    plot.caption = element_text(size = 5, margin = margin(t = 10), hjust = 0),
    legend.text.align = 0
  )

get_legend <- function(p) {
  tmp <- ggplotGrob(p)
  leg <- which(sapply(tmp$grobs, function(x) x$name) == "guide-box")
  tmp$grobs[[leg]]
}

legend <- get_legend(datasharing_plot)

datasharing_plot <- datasharing_plot + 
  coord_cartesian(ylim = c(0, 47)) + 
  theme(legend.position = "none")

response_plot <- response_plot + 
  coord_cartesian(ylim = c(0, 47)) + 
  theme(legend.position = "none") +
  ylab("") 

title_plot1 <- ggplot() + 
  labs(
    title = "Data sharing"
  ) +
  theme(
    plot.title = element_text(hjust = 0, size = 20, margin = margin(b = 0)),
    plot.margin = margin(l = 50, t = 5),
    panel.background = element_blank())

title_plot2 <- ggplot() + 
  labs(
    title = "Responses",
  ) +
  theme(
    plot.title = element_text(hjust = 0, size = 20, margin = margin(b = 0)),
    plot.margin = margin(l = 50, t = 5),
    panel.background = element_blank())

plot <- ggarrange(ggarrange(title_plot1, title_plot2, ncol = 2, labels = c("A", "B")),
                  legend,
                  ggarrange(datasharing_plot, response_plot, ncol = 2),
                  nrow = 3, heights = c(0.06,0.05, 1))


table_datasharing <- xtabs(count ~ dataavailab_per_study + DAS, data = dataavailab_das)
table_grob_datasharing <- tableGrob(table_datasharing, theme = ttheme_minimal(core = list(bg_params = list(fill = "transparent")), base_size = 15))

table_responses <- xtabs(count ~ response_per_study + DAS, data = responses_das_analysis)
table_grob_responses <- tableGrob(table_responses, theme = ttheme_minimal(core = list(bg_params = list(fill = "transparent")), base_size = 15))

combined_plot <- ggarrange(plot,
                           ggarrange(table_grob_datasharing, table_grob_responses, ncol = 2), 
                           nrow = 2, heights = c(1,0.1))

ggsave( combined_plot,
        filename = "Figure3.pdf",
        width = 14,  # Adjust the width of the image
        height = 10,  # Adjust the height of the image
        dpi = 1000,  # Set the minimum dpi
        units = "in"
)
```
