## Supplement 4: R Source code for analysis and creation of graphics for "A Case Study on Colposcopy-Based Cervical Cancer Staging Reveals an Alarming Lack of Data Sharing Hindering the Adoption of Machine Learning in Clinical Practice": README.txt

Related source code to article 'A Case Study on Colposcopy-Based Cervical Cancer Staging Reveals an Alarming Lack of Data Sharing Hindering the Adoption of Machine Learning in Clinical Practice'

## DESCRIPTION

This zip-File contains the related source code to reproduce results and graphics (please note that figure 2 was created using Word, hence no source code for creating figure 2 is included) using the Software R[1].

For the results of each section/subsection, an R-file is provided. Figure 1 and 3 can be reproduced via the file 'graphics.R'.

## USAGE

To execute the code, please set an appropriate working directory at the beginning of the R-file in which the Excel spreadsheet is stored (Supplement 3).

The following R-version was used: R version 4.3.2

The following R-packages are loaded to execute the code:
- gridExtra (Version 2.3)
- ggpubr (Version 0.6.0)
- tidyr (Version 1.3.0)    
- tidyverse (Version 2.0.0)
- dplyr (Version 1.1.4)
- readxl (Version 1.4.3)    
- ggplot2 (Version 3.4.4) 

Please see ## SESSION INFO for more information on the required packages and their versions.

## AUTHORS

Maxi Schulz[*], Andreas Leha[*]

[*] Department of Medical Statistics, University Medical Center Göttingen, Germany


## LICENSE

Copyright (c) 2024 Maxi Schulz

Permission is hereby granted, free of charge, to any person obtaining a copy of this software and associated documentation files (the “Software”), to deal in the Software without restriction, including without limitation the rights to use, copy, modify, merge, publish, distribute, sublicense, and/or sell copies of the Software, and to permit persons to whom the Software is furnished to do so, subject to the following conditions:

The above copyright notice and this permission notice shall be included in all copies or substantial portions of the Software.

THE SOFTWARE IS PROVIDED “AS IS”, WITHOUT WARRANTY OF ANY KIND, EXPRESS OR IMPLIED, INCLUDING BUT NOT LIMITED TO THE WARRANTIES OF MERCHANTABILITY, FITNESS FOR A PARTICULAR PURPOSE AND NONINFRINGEMENT. IN NO EVENT SHALL THE AUTHORS OR COPYRIGHT HOLDERS BE LIABLE FOR ANY CLAIM, DAMAGES OR OTHER LIABILITY, WHETHER IN AN ACTION OF CONTRACT, TORT OR OTHERWISE, ARISING FROM, OUT OF OR IN CONNECTION WITH THE SOFTWARE OR THE USE OR OTHER DEALINGS IN THE SOFTWARE.

## SESSION INFO

- Session info --------------------------------------------------------------------------------------------------------------
 setting  value
 version  R version 4.3.2 (2023-10-31 ucrt)
 os       Windows 10 x64 (build 17763)
 system   x86_64, mingw32
 ui       RStudio
 language (EN)
 collate  German_Germany.1252
 ctype    German_Germany.1252
 tz       Europe/Berlin
 date     2024-11-18
 rstudio  2023.06.0+421 Mountain Hydrangea (desktop)
 pandoc   NA

- Packages ------------------------------------------------------------------------------------------------------------------
 package     * version date (UTC) lib source
 abind         1.4-5   2016-07-21 [1] CRAN (R 4.3.1)
 backports     1.4.1   2021-12-13 [1] CRAN (R 4.3.1)
 broom         1.0.5   2023-06-09 [1] CRAN (R 4.3.2)
 cachem        1.0.8   2023-05-01 [1] CRAN (R 4.3.2)
 car           3.1-2   2023-03-30 [1] CRAN (R 4.3.2)
 carData       3.0-5   2022-01-06 [1] CRAN (R 4.3.2)
 cellranger    1.1.0   2016-07-27 [1] CRAN (R 4.3.2)
 cli           3.6.2   2023-12-11 [1] CRAN (R 4.3.2)
 colorspace    2.1-0   2023-01-23 [1] CRAN (R 4.3.1)
 cowplot       1.1.3   2024-01-22 [1] CRAN (R 4.3.2)
 devtools      2.4.5   2022-10-11 [1] CRAN (R 4.3.2)
 digest        0.6.33  2023-07-07 [1] CRAN (R 4.3.2)
 dplyr       * 1.1.4   2023-11-17 [1] CRAN (R 4.3.2)
 ellipsis      0.3.2   2021-04-29 [1] CRAN (R 4.3.2)
 fansi         1.0.6   2023-12-08 [1] CRAN (R 4.3.2)
 farver        2.1.1   2022-07-06 [1] CRAN (R 4.3.1)
 fastmap       1.1.1   2023-02-24 [1] CRAN (R 4.3.2)
 forcats     * 1.0.0   2023-01-29 [1] CRAN (R 4.3.1)
 fs            1.6.4   2024-04-25 [1] CRAN (R 4.3.3)
 generics      0.1.3   2022-07-05 [1] CRAN (R 4.3.1)
 ggplot2     * 3.4.4   2023-10-12 [1] CRAN (R 4.3.1)
 ggpubr      * 0.6.0   2023-02-10 [1] CRAN (R 4.3.3)
 ggsignif      0.6.4   2022-10-13 [1] CRAN (R 4.3.2)
 glue          1.6.2   2022-02-24 [1] CRAN (R 4.3.1)
 gridExtra   * 2.3     2017-09-09 [1] CRAN (R 4.3.2)
 gtable        0.3.4   2023-08-21 [1] CRAN (R 4.3.1)
 hms           1.1.3   2023-03-21 [1] CRAN (R 4.3.1)
 htmltools     0.5.7   2023-11-03 [1] CRAN (R 4.3.2)
 htmlwidgets   1.6.4   2023-12-06 [1] CRAN (R 4.3.2)
 httpuv        1.6.13  2023-12-06 [1] CRAN (R 4.3.2)
 labeling      0.4.3   2023-08-29 [1] CRAN (R 4.3.1)
 later         1.3.2   2023-12-06 [1] CRAN (R 4.3.2)
 lifecycle     1.0.4   2023-11-07 [1] CRAN (R 4.3.2)
 lubridate   * 1.9.3   2023-09-27 [1] CRAN (R 4.3.2)
 magrittr      2.0.3   2022-03-30 [1] CRAN (R 4.3.1)
 memoise       2.0.1   2021-11-26 [1] CRAN (R 4.3.2)
 mime          0.12    2021-09-28 [1] CRAN (R 4.3.1)
 miniUI        0.1.1.1 2018-05-18 [1] CRAN (R 4.3.2)
 munsell       0.5.0   2018-06-12 [1] CRAN (R 4.3.1)
 pillar        1.9.0   2023-03-22 [1] CRAN (R 4.3.1)
 pkgbuild      1.4.3   2023-12-10 [1] CRAN (R 4.3.2)
 pkgconfig     2.0.3   2019-09-22 [1] CRAN (R 4.3.1)
 pkgload       1.3.3   2023-09-22 [1] CRAN (R 4.3.2)
 profvis       0.3.8   2023-05-02 [1] CRAN (R 4.3.2)
 promises      1.2.1   2023-08-10 [1] CRAN (R 4.3.2)
 purrr       * 1.0.2   2023-08-10 [1] CRAN (R 4.3.1)
 R6            2.5.1   2021-08-19 [1] CRAN (R 4.3.1)
 ragg          1.2.7   2023-12-11 [1] CRAN (R 4.3.2)
 Rcpp          1.0.11  2023-07-06 [1] CRAN (R 4.3.1)
 readr       * 2.1.4   2023-02-10 [1] CRAN (R 4.3.2)
 readxl      * 1.4.3   2023-07-06 [1] CRAN (R 4.3.2)
 remotes       2.4.2.1 2023-07-18 [1] CRAN (R 4.3.2)
 rlang         1.1.2   2023-11-04 [1] CRAN (R 4.3.2)
 rstatix       0.7.2   2023-02-01 [1] CRAN (R 4.3.2)
 rstudioapi    0.15.0  2023-07-07 [1] CRAN (R 4.3.2)
 scales        1.3.0   2023-11-28 [1] CRAN (R 4.3.2)
 sessioninfo   1.2.2   2021-12-06 [1] CRAN (R 4.3.2)
 shiny         1.8.0   2023-11-17 [1] CRAN (R 4.3.2)
 stringi       1.8.3   2023-12-11 [1] CRAN (R 4.3.2)
 stringr     * 1.5.1   2023-11-14 [1] CRAN (R 4.3.2)
 systemfonts   1.0.5   2023-10-09 [1] CRAN (R 4.3.2)
 textshaping   0.3.7   2023-10-09 [1] CRAN (R 4.3.2)
 tibble      * 3.2.1   2023-03-20 [1] CRAN (R 4.3.1)
 tidyr       * 1.3.0   2023-01-24 [1] CRAN (R 4.3.1)
 tidyselect    1.2.0   2022-10-10 [1] CRAN (R 4.3.1)
 tidyverse   * 2.0.0   2023-02-22 [1] CRAN (R 4.3.2)
 timechange    0.3.0   2024-01-18 [1] CRAN (R 4.3.2)
 tzdb          0.4.0   2023-05-12 [1] CRAN (R 4.3.2)
 urlchecker    1.0.1   2021-11-30 [1] CRAN (R 4.3.2)
 usethis       2.2.2   2023-07-06 [1] CRAN (R 4.3.2)
 utf8          1.2.4   2023-10-22 [1] CRAN (R 4.3.1)
 vctrs         0.6.5   2023-12-01 [1] CRAN (R 4.3.2)
 withr         2.5.2   2023-10-30 [1] CRAN (R 4.3.1)
 xtable        1.8-4   2019-04-21 [1] CRAN (R 4.3.2)

## REFERENCES

[1] R Core Team (2021). R: A language and environment for statistical computing. R Foundation for Statistical Computing, Vienna, Austria. URL https://www.R-project.org/.
